# Autophagy gene expression in skeletal muscle of older individuals is associated with physical performance, muscle volume and mitochondrial function in the Study of Muscle, Mobility and Aging (SOMMA)

**DOI:** 10.1101/2023.11.04.23297979

**Authors:** Paul M Coen, Zhiguang Huo, Gregory J Tranah, Haley N Barnes, Peggy M Cawthon, Russell T Hepple, Frederico G S Toledo, Daniel S Evans, Olaya Santiago Fernández, Ana Maria Cuervo, Steven B Kritchevsky, Anne B Newman, Steven R Cummings, Karyn A Esser

**Affiliations:** California Pacific Medical Center Research Institute, San Francisco, California, USA; Department of Epidemiology and Biostatistics, University of California San Francisco, San Francisco, California, USA; Translational Research Institute, Advent Health, Orlando, Florida, USA; Department of Biostatistics, College of Public Health & Health Professions, College of Medicine University of Florida, Gainesville, Florida, USA; Department of Physical Therapy, University of Florida, Gainesville, Florida, USA; Department of Developmental & Molecular Biology, Albert Einstein College of Medicine, New York, New York, USA; Department of Internal Medicine, Wake Forest School of Medicine, Winston-Salem, NC, USA; Department of Epidemiology, School of Public Health, University of Pittsburgh, Pittsburgh, PA, USA; Department of Physiology and Ageing, College of Medicine, University of Florida, Gainesville, FL, USA; Department of Medicine, Division of Endocrinology and Metabolism, University of Pittsburgh School of Medicine, Pittsburgh, PA, USA

**Keywords:** mobility, aging, mitochondria, mTor, Autophagy, gene expression, oxidative metabolism

## Abstract

Autophagy is an essential component of proteostasis and a key pathway in aging. Identifying associations between autophagy gene expression patterns in skeletal muscle and physical performance outcomes would further our knowledge of mechanisms related with proteostasis and healthy aging. Muscle biopsies were obtained from participants in the Study of Muscle, Mobility and Aging (SOMMA). For 575 participants, RNA was sequenced and expression of 281 genes related to autophagy regulation, mitophagy and mTOR/upstream pathways were determined. Associations between gene expression and outcomes including mitochondrial respiration in muscle fiber bundles (MAX OXPHOS), physical performance (VO_2_ peak, 400m walking speed, and leg power), and thigh muscle volume were determined using negative binomial regression models. For autophagy, key transcriptional regulators including TFE3 and NFKB-related genes (RELA, RELB, NFKB1) were negatively associated with outcomes. On the contrary, regulators of oxidative metabolism that also promote overall autophagy, mitophagy and pexophagy (PPARGC1A, PPARA, EPAS1) were positively associated with multiple outcomes. In line with this, several mitophagy, fusion and fission related genes (NIPSNAP2, DNM1L, OPA1) were also positively associated with outcomes. For mTOR pathway and related genes, expression of WDR59 and WDR24, both subunits of GATOR2 complex (an indirect inhibitor of mTORC1) and PRKAG3, which is a regulatory subunit of AMPK, were negatively correlated with multiple outcomes. Our study identifies autophagy and selective autophagy such as mitophagy gene expression patterns in human skeletal muscle related to physical performance, muscle volume and mitochondrial function in older persons which may lead to target identification to preserve mobility and independence.

## INTRODUCTION

Aging is defined by a gradual loss of physiological integrity, with accrual of cellular damage widely considered the general cause. Several molecular hallmarks of aging have been proposed that contribute to accumulation of cellular damage, including genomic instability, loss of proteostasis, macroautophagy dysregulation, DNA damage and mitochondrial dysfunction (Lopez-Otin *et al*. 2023). In skeletal muscle, aging results in a loss of muscle mass, strength and oxidative capacity that contributes to lower cardiorespiratory fitness, slower walking speed and ultimately mobility limitations. These changes in muscle are linked to impaired protein homeostasis or proteostasis (Powers *et al*. 2009) including the loss of skeletal muscle proteins (Rooyackers *et al*. 1996; Balagopal *et al*. 1997; Short *et al*. 2005; Drummond *et al*. 2009), and alterations in the balance between protein synthesis and degradation (Rooyackers *et al*. 1996; Rasmussen *et al*. 2006). The molecular basis for these changes, including changes in the expression of several key genes and proteins implicated in growth, atrophy, proteosome degradation and mitochondrial metabolism, is also documented (Sandri 2002; Giresi *et al*. 2005; Short *et al*. 2005; Masiero *et al*. 2009; Sandri 2010).

Autophagy is the cellular process of lysosomal degradation and recycling of cytoplasmic components to maintain cellular homeostasis. Autophagy clears and recycles damaged organelles and macromolecules and preserves DNA stability (Sandri 2010; Schneider & Cuervo 2014; Kaushik *et al*. 2021). Autophagy decreases with aging in several tissues, however its role in regulating muscle mass and function remains poorly understood, particularly in humans. On one hand, excessive activation of autophagy aggravates muscle wasting (Mammucari *et al*. 2007) by removing portions of cytoplasm, proteins, and organelles.

Conversely, it was demonstrated that inhibition of autophagy can result in muscle degeneration and weakness (Masiero *et al*. 2009). Blocking autophagy in muscle can also lead to impacts on innervation, mitochondrial function, and oxidative damage (Carnio *et al*. 2014; Franco-Romero & Sandri 2021).

Autophagy is highly coordinated and is positively regulated by the energy sensor, AMP-activated protein kinase (AMPK) pathway, and negatively regulated by the nutrient sensor, the mammalian target of rapamycin (mTOR) pathway (Jung *et al*. 2010; Egan *et al*. 2011). The transcription factors, TFEB and TFE3, are recognized for their role regulating a network of genes (the CLEAR network) involved in lysosomal biogenesis, autophagy and lysosomal exocytosis (Sardiello *et al*. 2009). The nuclear localization of these factors is modulated by phosphorylation via both AMPK and mTORC pathways. The study of autophagy has led to the identification of multiple autophagy sub-types that selectively degrade organelles including mitophagy, the process of mitochondrial degradation, and pexophagy, which is the targeted degradation of peroxisomes. PGC-1α is a regulator of mitochondrial biogenesis and has been shown to also activate mitochondria fusion-fission events (Martin *et al*. 2014), autophagy and in particular mitophagy (Vainshtein *et al*. 2015; Yeo *et al*. 2019). While the extensive study of the regulation of autophagic flux (the combined process of autophagosome formation and clearance) in model systems has revealed this regulation to be complex and multifactorial, the relevant pathways that regulate autophagy and mitophagy in human muscle aging have yet to be fully elucidated.

To further understand the gene expression profiles related to autophagy in human muscle and how they associate with muscle and physical function phenotypes, we performed sequencing of RNA obtained from muscle biopsies collected from participants in the Study of Muscle, Mobility and Aging (SOMMA). SOMMA is a prospective, longitudinal study of older people at risk of major mobility disability designed to understand the contributions of skeletal muscle mass and key properties of muscle tissue from biopsies to mobility. Decreases in autophagy occur with aging and a few small studies have correlated protein markers of autophagy with compromised muscle function (Zeng *et al*. 2018; Aas *et al*. 2019). In this investigation, we examined associations between expression levels of 281 genes involved in autophagy with muscle mitochondrial function, 400-m walking speed, VO_2_ peak, leg strength, and thigh muscle volume. Here, we took a targeted candidate gene approach and hypothesized that expression of autophagy genes in the muscle of older persons are generally associated with muscle mitochondrial function and tissues beyond muscle, thereby substantially contributing to overall fitness, walking speed, strength, and muscle mass.

## RESULTS

### Participant characteristics

A total of 879 participants provided consent and completed baseline measurements across both clinical sites (Figure 1). Of the 879 participants with complete baseline measures, 591 participants had RNA sequencing completed and 575 of these had high quality sequencing and complete covariate data. The characteristics of the study population with complete RNA sequencing and complete covariate data are presented in Table 1.

**Figure 1.**
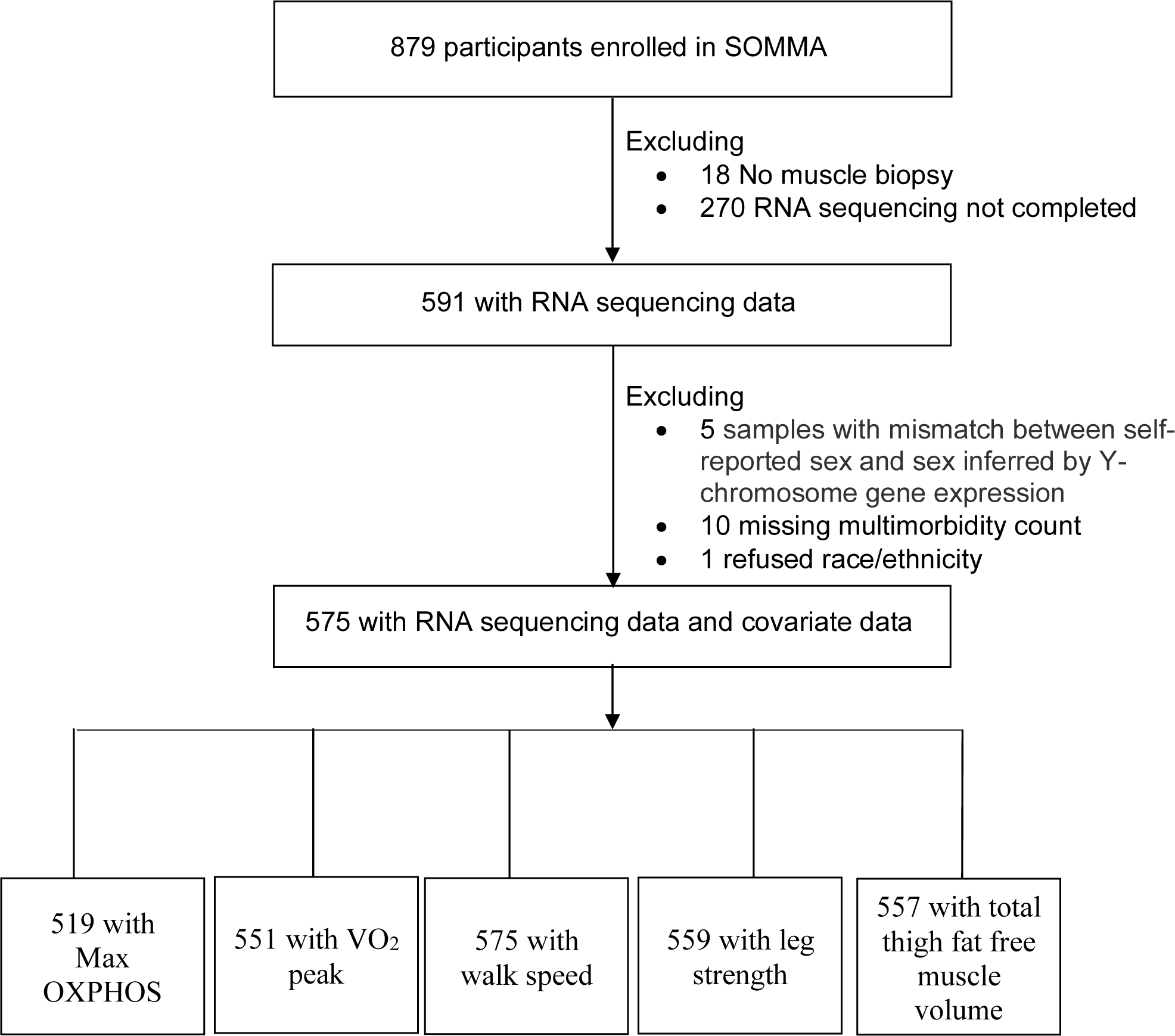
Participant inclusions. SOMMA participants selected for cross-sectional analyses.

**Table 1.**
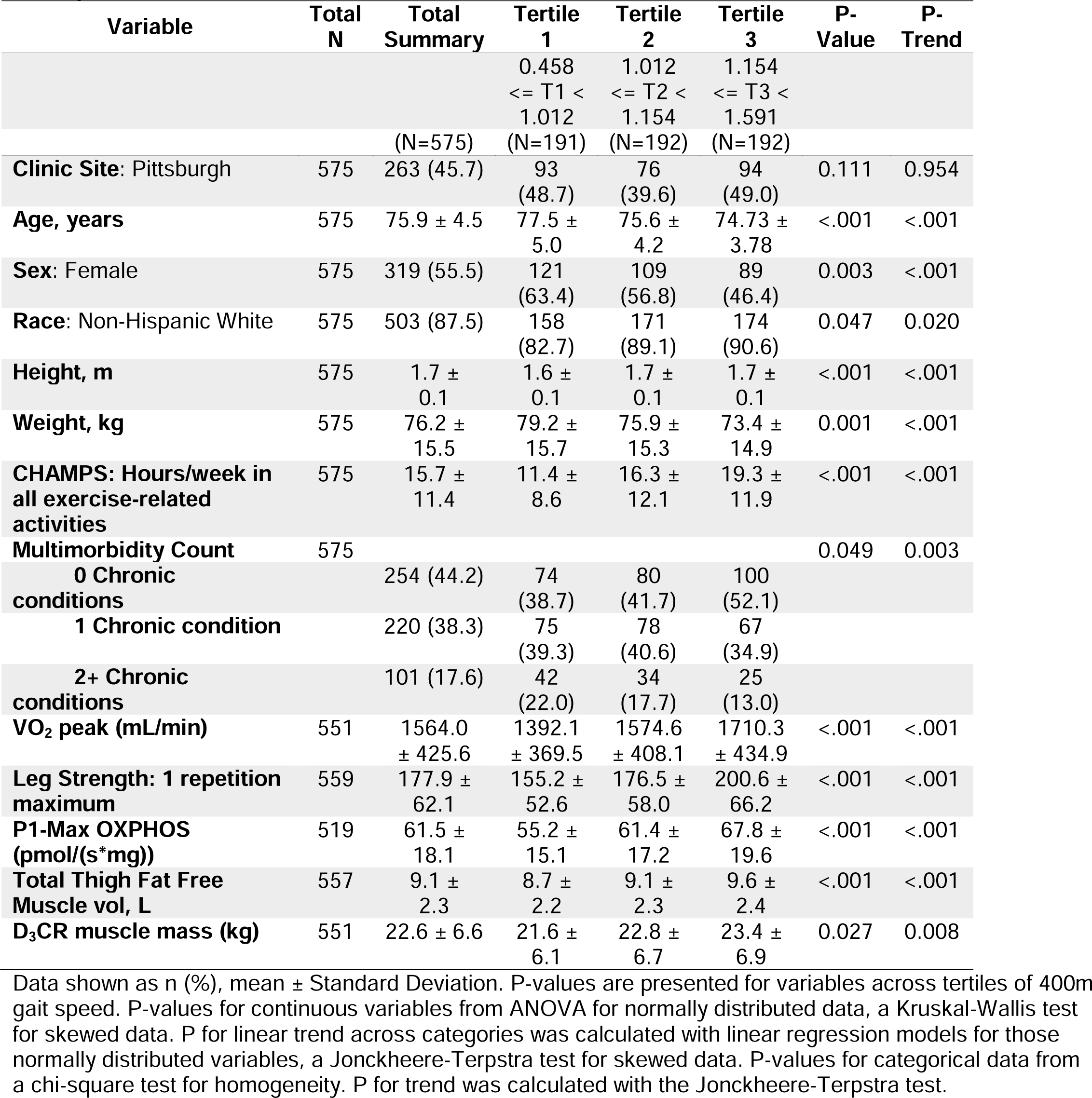
Baseline characteristics of included SOMMA participants also stratified by tertiles of 400m Walk Speed. Data shown as n (%), mean +/- Standard Deviation. P-values are presented for variables across tertiles of P1-Max OXPHOS. P-values for continuous variables from ANOVA for normally distributed data, a Kruskal-Wallis test for skewed data. P for linear trend across categories was calculated with linear regression models for those normally distributed variables, a Jonckheere-Terpstra test for skewed data. P-values for categorical data from a chi-square test for homogeneity. P for trend was calculated with the Jonckheere-Terpstra test.

| Variable | Total N | Total Summary | Tertile 1 | Tertile 2 | Tertile 3 | P-Value | P-Trend |
| --- | --- | --- | --- | --- | --- | --- | --- |
|  |  |  | 0.458<br>≤ T1 <<br>1.012 | 1.012<br>≤ T2 <<br>1.154 | 1.154<br>≤ T3 <<br>1.591 |  |  |
|  |  | (N=575) | (N=191) | (N=192) | (N=192) |  |  |
| **Clinic Site: Pittsburgh** | 575 | 263 (45.7) | 93<br>(48.7) | 76<br>(39.6) | 94<br>(49.0) | 0.111 | 0.954 |
| **Age, years** | 575 | 75.9 ± 4.5 | 77.5 ± 5.0 | 75.6 ± 4.2 | 74.73 ± 3.78 | <.001 | <.001 |
| **Sex: Female** | 575 | 319 (55.5) | 121<br>(63.4) | 109<br>(56.8) | 89<br>(46.4) | 0.003 | <.001 |
| **Race: Non-Hispanic White** | 575 | 503 (87.5) | 158<br>(82.7) | 171<br>(89.1) | 174<br>(90.6) | 0.047 | 0.020 |
| **Height, m** | 575 | 1.7 ± 0.1 | 1.6 ± 0.1 | 1.7 ± 0.1 | 1.7 ± 0.1 | <.001 | <.001 |
| **Weight, kg** | 575 | 76.2 ± 15.5 | 79.2 ± 15.7 | 75.9 ± 15.3 | 73.4 ± 14.9 | 0.001 | <.001 |
| **CHAMPS: Hours/week in all exercise-related activities** | 575 | 15.7 ± 11.4 | 11.4 ± 8.6 | 16.3 ± 12.1 | 19.3 ± 11.9 | <.001 | <.001 |
| **Multimorbidity Count** | 575 |  |  |  |  | 0.049 | 0.003 |
| **0 Chronic conditions** |  | 254 (44.2) | 74<br>(38.7) | 80<br>(41.7) | 100<br>(52.1) |  |  |
| **1 Chronic condition** |  | 220 (38.3) | 75<br>(39.3) | 78<br>(40.6) | 67<br>(34.9) |  |  |
| **2+ Chronic conditions** |  | 101 (17.6) | 42<br>(22.0) | 34<br>(17.7) | 25<br>(13.0) |  |  |
| **VO<sub>2</sub> peak (mL/min)** | 551 | 1564.0 ± 425.6 | 1392.1 ± 369.5 | 1574.6 ± 408.1 | 1710.3 ± 434.9 | <.001 | <.001 |
| **Leg Strength: 1 repetition maximum** | 559 | 177.9 ± 62.1 | 155.2 ± 52.6 | 176.5 ± 58.0 | 200.6 ± 66.2 | <.001 | <.001 |
| **P1-Max OXPHOS (pmol/(s\*mg))** | 519 | 61.5 ± 18.1 | 55.2 ± 15.1 | 61.4 ± 17.2 | 67.8 ± 19.6 | <.001 | <.001 |
| **Total Thigh Fat Free Muscle vol, L** | 557 | 9.1 ± 2.3 | 8.7 ± 2.2 | 9.1 ± 2.3 | 9.6 ± 2.4 | <.001 | <.001 |
| **D<sub>3</sub>CR muscle mass (kg)** | 551 | 22.6 ± 6.6 | 21.6 ± 6.1 | 22.8 ± 6.7 | 23.4 ± 6.9 | 0.027 | 0.008 |

### RNA (human Ensembl genes (ENSG)) detection

The mean, median, and SD of the PCR duplicate percent per sample was 59%, 56% and 9%, respectively (Supplementary Table 1). After PCR duplicates were removed, the number of aligned reads per sample was high (mean=69,117,209, median = 71,313,059, SD = 14,444,848, range = 12,853,785-102,724,183).

### Association of autophagy gene expression with multiple outcomes

We utilized a published curated list of autophagy genes (Bordi *et al*. 2021) to conduct a targeted analysis of the RNASeq dataset. A total of 281 genes related to autophagy regulation (68 genes), mitophagy (80 genes), and mTOR and upstream pathways (133 genes), were analyzed (Supplemental table 2). For each gene set, we examined the genes that were most significantly associated across multiple outcomes (Figure 2). All results report log base 2-fold changes reflecting the change in gene expression per one SD unit increase in each trait.

**Figure 2.**
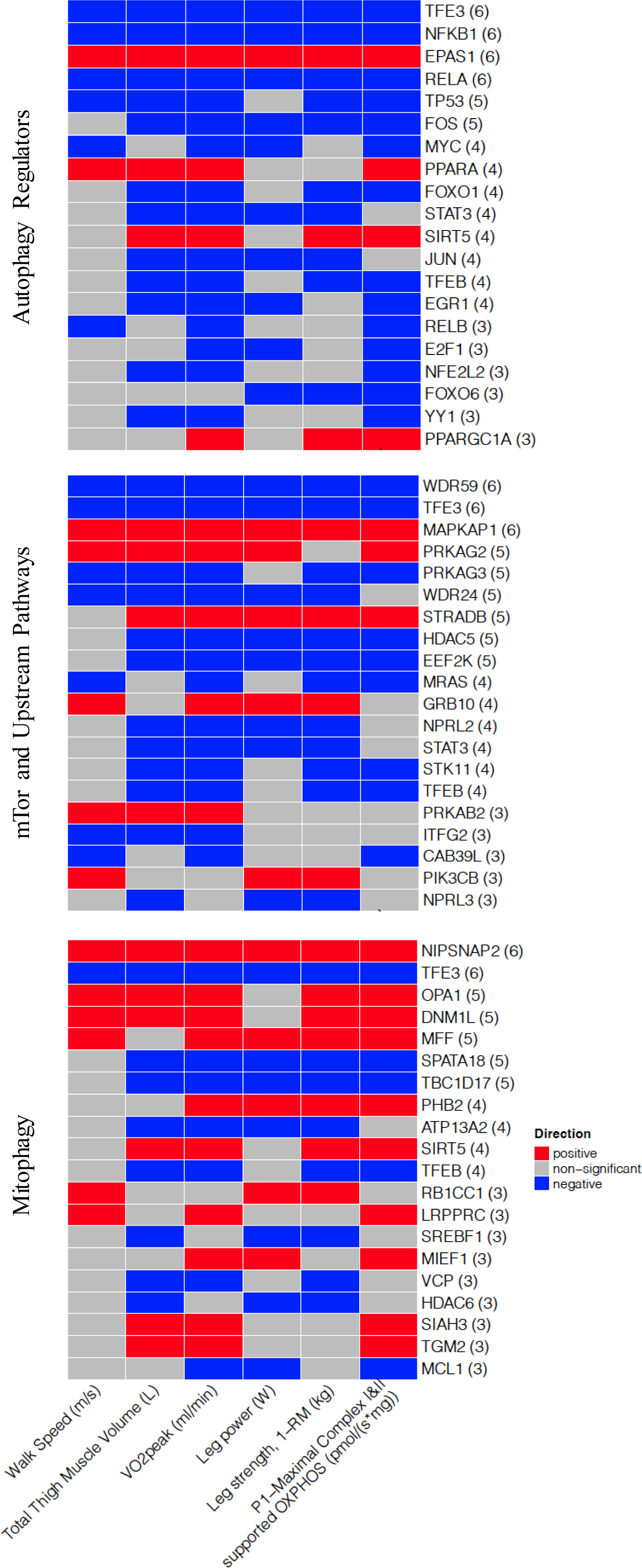
Significant associations of autophagy genes with mitochondrial function, physical performance, and muscle mass measures. Heat map capturing top 20 statistically significant (p < 0.05 FDR adjusted) genes identified by our models: each color represents positive (red) or negative (blue) associations.

Analysis of the curated list of autophagy genes did not reveal any significant correlations between the core autophagy machinery (Beclin, Atg7, Atg5) and outcomes (Max OXPHOS, VO_2_ peak). For autophagy regulators, key transcriptional regulators including TFE3, TFEB, NFKB related genes (RELA, RELB, NFKB1), FOS, and FOXO1 were all significantly negatively associated with associated with 400m walk speed, VO_2_ peak, and Max OXPHOS. Alternatively, regulators of oxidative metabolism that also promote autophagy and mitophagy including PPARGC1A, PPARA, and EPAS1, a driver of pexophagy(Schonenberger *et al*. 2015), were all positively associated with VO_2_ peak, and Max OXPHOS. We also found that several mitochondria, fusion and fission related genes (NIPSNAP2, DNM1L, OPA1) were also positively associated with multiple outcomes including VO_2_ peak, Max OXPHOS and 400m walking speed. For mTOR pathway and related genes, expression of WDR59 and WDR24, both subunits of GATOR2 complex (an inhibitor of mTORC1) and PRKAG3, which is a regulatory subunit of AMPK, were negatively associated with multiple outcomes including 400m walk speed, VO_2_ peak, leg strength, leg power and thigh muscle volume. In contrast, MAPKAP1, which is a subunit of mTORC2 was positively associated with outcomes. A summary of all statistically significant associations for each gene set: autophagy regulation, mitophagy, and mTOR and upstream pathways with each trait is presented in Supplemental tables 3-8.

### Association of mitochondrial respiration with gene expression

We next examined associations between gene expression for all three gene sets and specific outcomes. Maximal Complex I&II supported OXPHOS was positively associated with the expression of the deacetylases SIRT5, SIRT3, the regulators of mitochondrial dynamics MFN2, MUL1, and the component of the vacuolar proton pump ATPV1B1 (Figure 3). Conversely, OXPHOS was negatively associated with the transcription factors FOS, MYC and the regulator of protein translation EEF2K, among others. The full list of associations between autophagy genes and Max OXPHOS is presented in Supplementary Table 3.

**Figure 3.**
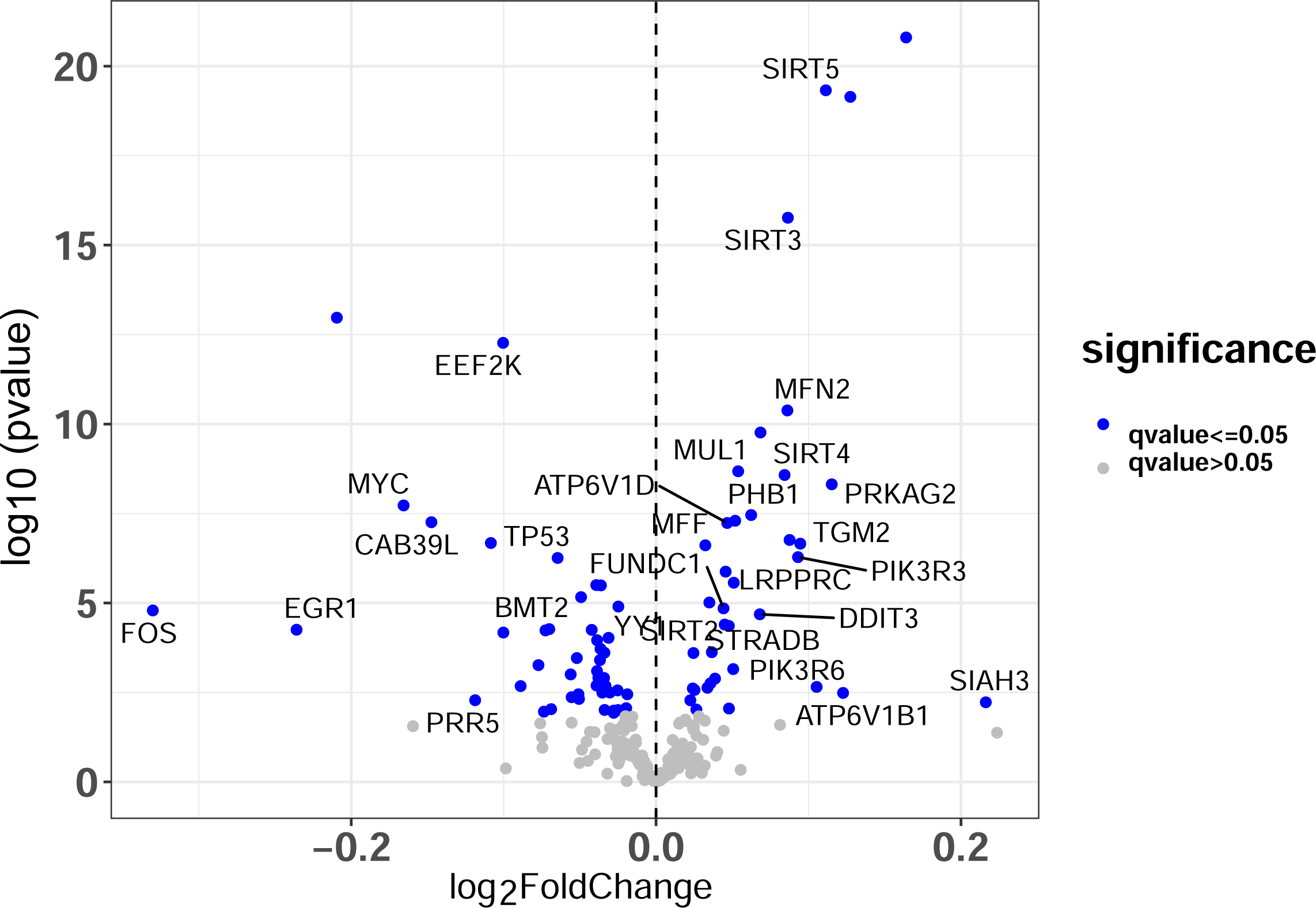
Associations with Max OXPHOS. Volcano plot capturing all statistically significant (p < 0.05 FDR adjusted) genes identified by our models: Each dot represents a gene; the dot color indicates significance level. Base model: gene expression∼ Max OXPHOS + age + gender + clinic site + race/ethnicity + height + weight + physical activities + multimorbidity count + sequencing batch.

### Association of VO_2_ peak with gene expression

Autophagy genes that were positively associated with higher VO_2_ peak, included the autophagy regulators PPARGC1A, PPARA, the mitochondria dynamic regulators related to mitophagy DNM1L and OPA1, the mitophagy core gene NIPSNAP2, and the pexophagy-related hypoxia induced gene EPAS1 (Figure 4). Genes that were negatively associated with VO_2_ peak, included the autophagy regulators RELA, RELB, and the regulators of AMPK and mTOR, PRKAG3 and WDR24, respectively. The full list of associations between autophagy genes and Max OXPHOS is presented in Supplementary Table 4.

**Figure 4.**
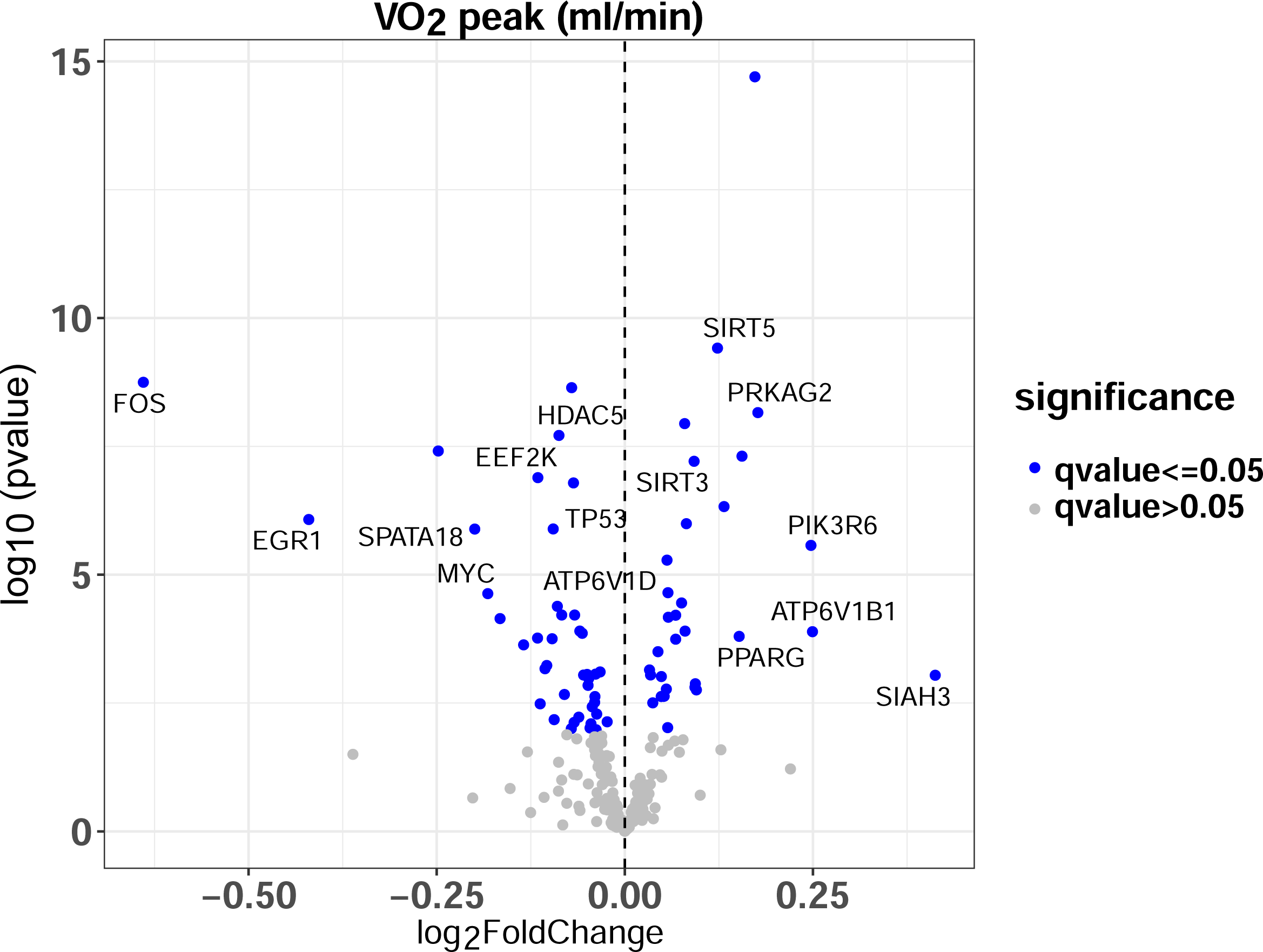
Associations with VO_2_ peak. Volcano plot capturing all statistically significant (p < 0.05 FDR adjusted) genes identified by our models: Each dot represents a gene; the dot color indicates significance level. Base model: gene expression∼ VO_2_ Peak + age + gender + clinic site + race/ethnicity + height + weight + physical activities + multimorbidity count + sequencing batch.

### Association of 400m walk speed with gene expression

Finally, we examined associations between autophagy-related gene expression and 400m walk speed. Genes that were positively associated with higher 400m walk speed, included mTOR and upstream pathway genes PRKAG2, PRKAB2, and TSC1 (Figure 5). Genes that were negatively associated with 400m walk speed, included positive (TFEC, TFE3) and negative (ATF5) autophagy regulators and the mTOR regulator SESN3. The full list of associations between autophagy genes and 400m walk speed is presented in Supplementary Table 5. The list of associations between autophagy genes and leg strength, leg power and thigh muscle volume are presented in Supplementary Tables 6-8.

**Figure 5.**
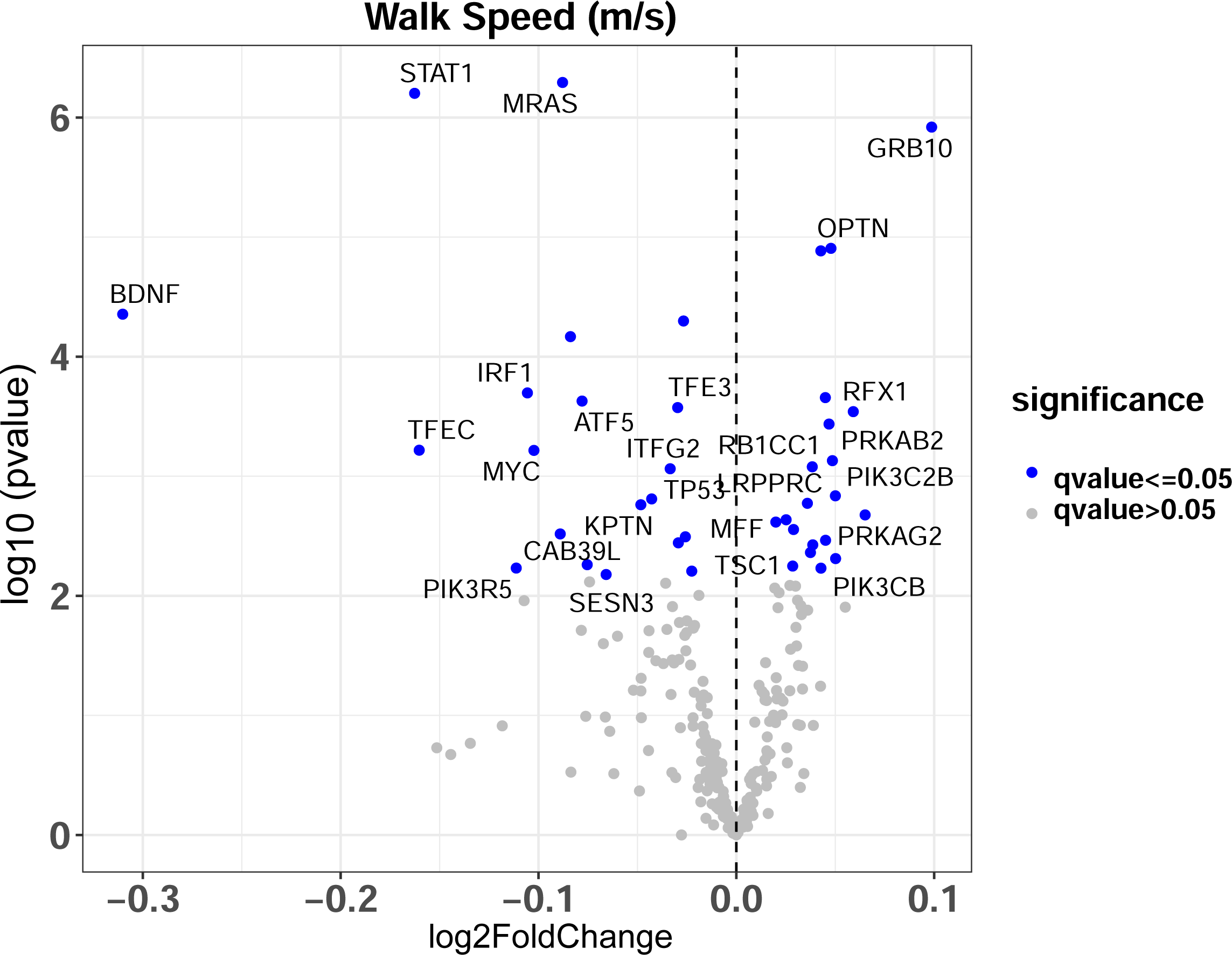
Associations with 400m Walk Speed. Volcano plot capturing all statistically significant (p < 0.05 FDR adjusted) genes identified by our models: Each dot represents a gene; the dot color indicates significance level. Base model: gene expression∼ walk speed + age + gender + clinic site + race/ethnicity + height + weight + physical activities + multimorbidity count + sequencing batch.

## DISCUSSION

Autophagy is a critical mechanism for maintaining cellular homeostasis, the dysregulation of which has been implicated as a key contributor to the biology of aging. Despite this, the relevance of skeletal muscle autophagy to measures of physical function, fitness and muscle mass and mitochondrial function in older adults has received relatively little attention. Here we leveraged transcriptomic data generated from muscle biopsy samples obtained from adults participating in the Study of Muscle, Mobility and Aging (SOMMA) to investigate the relationships between autophagy gene expression and multiple aging muscle and whole-body phenotypes (Cummings *et al*. 2023). We identified distinct patterns of association between expression of genes known to be key transcriptional regulators and signaling molecules of the autophagy program with several functional and physical domains. These data provide unique insight into the transcriptional regulation of autophagy in clinically relevant human aging phenotypes.

The principal finding was that several transcriptional regulators of the autophagy/lysosome pathway including TFE3, TFEB, FOS, and FOXO1 were all negatively associated with VO_2_ peak, Max OXPHOS, leg strength and thigh muscle volume. These data suggest that the downregulation of autophagy/lysosome transcriptional program is associated with better muscle and whole-body performance. However, because autophagy is part of the cellular response to several stressors including deprivation of nutrients or growth factors, and hypoxia and protects cellular homeostasis against oxidative stress and inflammation (Javali *et al*. 2023), lower expression of the autophagy/lysosomal transcriptional program could be a consequence of reduced overall cellular stress. Our results imply that older adults with higher fitness/respiration and walking speed may have lower levels of oxidative stress and inflammation and thus lower expression of drivers of the autophagy program. This finding is congruent with reports that older adults who engage in exercise and who are more fit have lower indices of inflammation including lower TNFα and IL-1β due to exercise-induced IL-6 release from muscle (Starkie *et al*. 2003; Pedersen & Fischer 2007). This data is also in-line with reports that higher levels of circulating inflammatory mediators are associated with poor mobility in older adults (Penninx *et al*. 2004). In further support of this possibility, we found modest or no correlation with expression of many of the core autophagy machinery, suggesting that it is mostly inducible autophagy (activated in response to stress) that is upregulated in adults with lower muscle fitness.

We found that expression of PPARA and PPARGC1A were positively associated with multiple outcomes including VO_2_ peak and Max OXPHOS. PPARGC1A, the gene that encodes PGC-1α, is most well-known for its role in mitochondrial biogenesis but has been reported to protect muscle mass in the context of muscle atrophy triggered by aging (Yang *et al*. 2020), chronic heart failure (Geng *et al*. 2011) and various other muscle wasting conditions (Sandri *et al*. 2006). More recently, it has been shown that PPARGC1A also plays a role in activating the autophagy-lysosome pathway (Takikita *et al*. 2010; Tsunemi *et al*. 2012) and specifically mitophagy in muscle (Vainshtein *et al*. 2015). In addition, PPARGC1A inhibits the expression of multiple pro-inflammatory mediators, including TNF-α and IL-6 (Handschin *et al*. 2007) and regulates antioxidant defense mechanisms (Kang & Li Ji 2012), both of which in turn can activate autophagy. In line with the role of PPARGC1A in mitophagy, we found that several mitochondria, fusion and fission related genes (DNM1L, OPA1) and the mitophagy “eat me” signal NIPSNAP2 were also positively associated with multiple outcomes, including VO_2_ peak, Max OXPHOS and 400m walking speed. In rodent models, deletion of the mitochondria fusion mediator OPA1 in skeletal muscle, caused mitochondrial dysfunction, oxidative stress and inflammation (Tezze *et al*. 2017; Rodriguez-Nuevo *et al*. 2018). DNM1L (or Drp-1) is a mediator of mitochondrial fission and muscle-specific Drp1 knockout in mice leads to a severe myopathic phenotype including muscle wasting and weakness (Favaro *et al*. 2019) and Drp1 knock down also decreases in mitochondrial respiration and increases markers of muscle denervation, fibrosis, and oxidative stress and also alters autophagy and mitophagy levels (Dulac *et al*. 2020). Taken together, these findings indicate that mitophagy and mitochondrial fusion and fission are essential for skeletal muscle health. However, studies in humans did not detect any impact of aging per se on Opa1 protein content in the skeletal muscle of sedentary young and old humans (Distefano *et al*. 2017), while physically active older adults had greater expression of Opa1 protein (Tezze *et al*. 2017). With respect to Drp1 expression, some studies report a decreased (Distefano *et al*. 2017) or an unchanged (Joseph *et al*. 2012) Drp1 content in skeletal muscle of older individuals, while physically active older adults had greater expression of Drp1 protein (Tezze *et al*. 2017). Based on these prior studies the consensus seems to be that mitochondrial dynamics do not play a role in muscle aging. However, unlike the previous study designs, our analysis focuses on associations between mitophagy gene expression and age-related phenotypes rather than aging *per se*.

mTORC1 is a master regulator of cell growth and metabolism through sensing and integrating different nutritional and environmental cues. Dysregulation and hyperactivation of mTOR contributes to several age-related diseases, such as cancer, neurodegenerative diseases, and type 2 diabetes mellitus. In skeletal muscle, mTORC1 signaling plays a critical role in protein synthesis and degradation, including regulating many steps of autophagy. The decline in skeletal muscle mass with aging is complex but anabolic resistance, or the limited ability to increase protein synthesis and inhibit autophagy in response to stimuli such as amino acids, is implicated as a contributing factor. In this study, we found that expression of mTOR pathway related genes WDR59 and WDR24 were both negatively correlated with 400m walk speed, VO_2_ peak, leg strength, leg power and thigh muscle volume. WDR59 and WDR24 are both subunits of GATOR2 complex which functions as a positive regulator of amino-acid-mediated mTORC1 activation (Bar-Peled *et al*. 2013) and WDR24 modulates mTOR through the previously described changes in lysosome cellular positioning and dynamics (Korolchuk *et al*. 2011), and thus indirectly modulates autophagic flux (Cai *et al*. 2016). This is in line with the observation that TFEB expression is also negatively associated with outcomes as mTORC1 regulates TFEB localization to the nucleus and activation by phosphorylating the transcription factor on several serine and threonine residues (Rabanal-Ruiz *et al*. 2017). Taken together, the associations of WDR59, WDR24 and TFEB with outcomes suggest that greater performance/fitness in older adults could be related to a blunted capacity of muscle mTORC1 to inhibit autophagy.

Our study has limitations. The analysis was performed on whole muscle tissue and the impact of mitochondrial content and muscle fiber type proportions on gene expression are not accounted for. For example, we observed that several genes whose expression correlated with Max OXPHOS are expressed exclusively within (or on) the mitochondria (SIRT5, SIRT3, MFN2, MUL1). That said, examining relationships between gene expression and relevant aging phenotypes in whole tissue initially is important to establish proof of concept/mechanism. A second limitation is that the study participants are of mostly White ancestry, thus potentially limiting generalizability of findings. Our data are cross-sectional and observational which limits our ability or prove causality. A strength of our study is that we paired muscle gene expression and mitochondrial function with several measures of fitness, strength, and muscle mass and accounted for potential confounding factors. Previous studies did not include older adults at risk of mobility disability and only analyzed cross-sectional associations between one or two properties and physical performance. Moreover, by focusing on a curated set of genes, rather than the broader gene ontology families, we were able to test specific hypotheses regarding the role of autophagy across mitochondrial function, fitness, mobility, strength, and muscle volume.

## CONCLUSION

This study of autophagy genes in 575 SOMMA participants has revealed significant associations between autophagy regulation, mitophagy and mTOR pathways genes and a diverse range of clinically-relevant phenotypes that include walking speed, VO_2_ peak, maximal mitochondrial respiration, total thigh muscle volume, leg power and leg strength. Our results support and add to the evidence that transcriptional regulation of autophagy plays an important role in driving declines in various indices of muscle function that are key to mobility with aging. Additional studies are needed to further decipher the role of autophagy, including the contribution of post-transcriptional regulation, in skeletal muscle fitness during aging.

## EXPERIMENTAL PROCEDURES

### Study population

The Study of Muscle, Mobility and Aging (SOMMA) is a prospective cohort study of mobility in community-dwelling older adults. Participants for the current study were from the baseline cohort, enrolled between April 2019 and December 2021 (Cummings *et al*. 2023). SOMMA was conducted at 2 clinical sites: University of Pittsburgh (Pittsburgh, PA) and Wake Forest University School of Medicine (Winston-Salem, NC). Eligible participants were ≥70 years old at enrollment, had a body mass index (BMI) of 18–40 kg/m^2^, and were eligible for magnetic resonance (MR) imaging and a muscle tissue biopsy (Cummings *et al*. 2023). Individuals were further excluded if they had active cancer or were in the advanced stages of heart failure, renal failure on dialysis, dementia, or Parkinson’s disease. Participants must have been able to complete the 400-meter walk; those who appeared as they might not be able to complete the 400m walk at the in-person screening visit completed a short distance walk (4 meters) to ensure their walking speed as >=0.6m/s. The study protocol was approved by the Western Institutional Review Board Copernicus Group (WCG IRB; study number 20180764) and all participants provided written informed consent. In brief, baseline testing occurred across 3 separate days of clinic visits that were generally within 6-8 weeks of each other. The mean time between Day 1 and 3 was 42 days or ∼ 6 weeks. Day 1 included general clinic assessments (e.g., physical and cognitive tests; 5 hours), Day 2 included magnetic resonance imaging and Cardiopulmonary Exercise Testing (MR and CPET, 2–3 hours), and Day 3 included fasting specimen and tissue collection (2 hours). There were 879 participants who completed Day 1 of baseline testing and had at least one primary SOMMA measure: CPET, MR imaging, or muscle tissue biopsy.

### Demographic, health, and functional measures

#### Cardiorespiratory fitness (VO_2_ peak)

Cardiorespiratory fitness was measured using gold standard VO_2_ peak (mL/min) from Cardiopulmonary Exercise Testing (CPET). A standardized CPET, using a modified Balke or manual protocol, was administered to participants to measure ventilatory gases, oxygen and carbon dioxide inhaled and exhaled during exercise (Balady *et al*. 2010). Two slow 5-minute walking tests were conducted before and after the maximal effort test to assess walking energetics at preferred walking speed and a slow fixed speed of 1.5 mph. Participants who were excluded from the maximal effort symptom-limited peak test had acute electrocardiogram (ECG) abnormalities, uncontrolled blood pressure or history of myocardial infarction, unstable angina or angioplasty in the preceding 6 months. Testing for VO_2_ peak began at the participant’s preferred walking speed with incremental rate (0.5 mph) and/or slope (2.5%) increased in 2-minute stages until respiratory exchange ratio, ratio between VCO_2_ and VO_2_, was ≥1.05 and self-reported Borg Rating of Perceived Exertion (Borg 1982) was ≥17. Blood pressure, pulse oximetry, and ECG were monitored throughout exercise. VO_2_ peak was determined in the BREEZESUITE software (MGC Diagnostics, St. Paul, MN) as the highest 30-second average of VO2 (L/min) achieved. The data were manually reviewed to ensure the correct VO_2_ peak was selected for each participant.

#### Other measures

Participants were asked to walk at their usual pace for 400 m from which walking speed (m/s) was calculated. Whole-body D_3_Cr muscle mass was measured in participants using a d3-creatine dilution protocol as previously described (Stimpson *et al*. 2012; Stimpson *et al*. 2013). Knee extensor leg power was assessed using a Keiser Air 420 exercise machine in the same leg as the muscle biopsy. Resistance to test power was based on determination of the 1 repetition maximum leg extensor strength. Weight was assessed by balance beam or digital scales and height by wall-mounted stadiometers. An approximately 6-minute-long MR scan was taken of the whole body to assess body composition including thigh muscle volume with image processing by AMRA Medical (Linge *et al*. 2018). The CHAMPS questionnaire (Stewart *et al*. 2001) was used to assess specific types and the context of physical activities. Participants were asked to self-report physician diagnosis of cancer (excluding nonmelanoma skin cancer), cardiac arrythmia, chronic kidney disease, chronic obstructive pulmonary disease, coronary artery disease, congestive heart failure, depression, diabetes, stroke, and aortic stenosis; from this list a multimorbidity count (0, 1, or 2+) was calculated.

#### Body Size

Weight was assessed by balance beam or digital scales and height by wall-mounted stadiometers.

### Gene expression and mitochondrial respiration measurements

#### Skeletal muscle biopsy collection and processing

Percutaneous biopsies were collected from the middle region of the musculus vastus lateralis under local anesthesia using a Bergstrom canula with suction. The specimen was blotted dry of blood and interstitial fluid and dissected free of any connective tissue and intermuscular fat. Approximately 20 mg of the biopsy specimen was placed into ice-cold BIOPS media (10 mM Ca–EGTA buffer, 0.1 M free calcium, 20 mM imidazole, 20 mM taurine, 50 mM potassium 2-[N-morpholino]-ethanesulfonic acid, 0.5 mM dithiothreitol, 6.56 mM MgCl2, 5.77 mM ATP, and 15 mM phosphocreatine [PCr], pH 7.1) for respirometry, as previously described (Mau *et al*. 2023). Myofiber bundles of approximately 2–3 mg were teased apart using a pair of sharp tweezers and a small Petri dish containing ice-cold BIOPS media. After mechanical preparation, myofiber bundles were chemically permeabilized for 30 minutes with saponin (2 mL of 50 μg/mL saponin in ice-cold BIOPS solution) placed on ice and a rocker (25 rpm). Myofiber bundles were washed twice (10 min each) with ice-cold MiR05 media (0.5 mM ethylenediaminetetraacetic acid, 3 mM MgCl2·6H2O, 60 mM K-lactobionate, 20 mM taurine, 10 mM KH2PO4, 20 mM N-2-hydroxyethylpiperazine-NL-2-ethanesulfonic acid, 110 mM sucrose, and 1 g/L bovine serum albumin, pH 7.1) on an orbital shaker (25 rpm). The second wash in MiR05 contained blebbistatin (25 μM), a myosin II ATPase inhibitor, that was used to inhibit muscle contraction. Fiber bundle wet weight was determined immediately after permeabilization using an analytical balance (Mettler Toledo, Columbus, OH).

#### Mitochondrial respiration

Maximal complex I- and II-supported oxidative phosphorylation (P1-Max OXPHOS, also known as State 3 respiration) was measured in permeabilized muscle fiber bundles from biopsies as previously described (Mau *et al*. 2023).

#### RNA Library preparation and sequencing

Total RNA from frozen human skeletal muscle samples (∼5 to 30mg) was prepared using Trizol solution (Invitrogen) according to manufacturer’s direction in 2.0mL Eppendorf safe-lock tubes. Homogenization was performed using the Bullet Blender (NextAdvance, Raymertown NY USA) with an appropriate quantity of stainless-steel beads (autoclaved, 0.5∼2mm, NextAdvance, Raymertown NY USA) at 4°C on Setting 8 (Max is 12) in 30 second bouts. The homogenization step was repeated 5 times for a total of 3 minutes with at least 1 minute break between each bout. The removal of residual genomic DNA was performed by incubating the RNA sample with DNase (AM1907, Thermosci) plus RiboLock RNase inhibitor (EO0381, Thermisci) at 37°C for 25min in a heating block (400rpm). Cleanup of the RNA samples was done using the DNase inactivation reagent following instructions provided by the manufacturer (AM1907, Thermosci). The RNA concentration and integrity were determined by using ThermoSci Nanodrop and Agilent Tapestation.

To prepare RNAseq library, polyA mRNA was isolated from about 250ng total RNA by using NEBNext Poly(A) mRNA magnetic isolation module (E7490L, NEB) and mRNA library constructed by using NEBNext Ultra II directional RNA library Pre Kit for Illumina (E7760L, NEB). Equal molarity of RNAseq libraries were pooled and sequenced on Illumina NovaSeq (2X150bp) to reach 80M reads per sample.

#### Alignment and quality control

The reads from RNA-sequencing were aligned to the Genome Reference Consortium Human Build 38 (GRCh38) using the HISAT2 software(Kim *et al*. 2019). Duplicated aligned reads were further marked and excluded using the Picardtools software (http://broadinstitute.github.io/picard/). Expression count data were obtained using the HTseq software (Anders *et al*. 2015). Genes with a total count of ≤ 20 across all samples were filtered out to remove non-expressed genes. The quality of the RNASeq data was gauged by the alignment rate and the duplication rate.

#### Association analyses

Expression levels of 281 genes related to mTOR and upstream pathways (133 genes), autophagy regulation (68 genes), and selective autophagy of mitochondria (mitophagy) (80 genes) were analyzed (Bordi *et al*. 2021). Gene expression associations with traits were identified using negative binomial regression models as implemented by DESeq2 (Love *et al*. 2014) in R and adjusted for age, gender, clinic site, race/ethnicity, height, weight, hours per week in all exercise-related activities (CHAMPS), multimorbidity count category, and sequencing batch. DESeq2 uses a negative binomial generalized linear model for differential analysis and applies the Wald test for the significance of GLM coefficients. The Benjamini-Hochberg false discovery rate method was used for P-value adjustment. Genes were considered differentially expressed according to the significance criteria of FDR<0.05. In negative binomial models, traits were modeled using the number of standard deviations (SDs) from each trait’s mean. Consequently, the reported log base 2-fold changes reflect the change in gene expression per one SD unit increase in each trait. Volcano plots were created to visualize the differential expression of RNAs (ENSGs) associated with functional measures. Heat maps were created to summarize significant ENSG associations across all analyzed traits.

## Statement relating to relevant ethics and integrity policies

The study protocol was approved by the Western Institutional Review Board Copernicus Group (WCG IRB; study number 20180764) and all participants provided written informed consent.

## Acknowledgments

We acknowledge all SOMMA staff and investigators, and we thank all the SOMMA participants who enabled this research. For a full list of personnel who contributed to the SOMMA study, please see Cummings SR, Newman AB, Coen PM, Hepple RT, Collins R, Kennedy K, Danielson M, Peters K, Blackwell T, Johnson E, Mau T, Shankland EG, Lui LY, Patel S, Young D, Glynn NW, Strotmeyer ES, Esser KA, Marcinek DJ, Goodpaster BH, Kritchevsky S, Cawthon PM. The Study of Muscle, Mobility and Aging (SOMMA). A Unique Cohort Study about the Cellular Biology of Aging and Age-related Loss of Mobility. J Gerontol A Biol Sci Med Sci. 2023 Feb 9:glad052. doi: 10.1093/gerona/glad052. Epub ahead of print. PMID: 36754371.

## Conflict of Interest statement

S.R.C. is a consultant to Bioage Labs. P.M.Ca. is a consultant to and owns stock in MyoCorps. All other authors declare no conflict of interest. A.M.C is a consultant to Generian Therapeuthics and it is a consultant to and owns stock in Life Biosciences.

## Funding statement

SOMMA is funded by the National Institute on Aging (NIA) grant number R01AG059416. Study infrastructure support was funded in part by NIA Claude D. Pepper Older American Independence Centers at University of Pittsburgh (P30AG024827) and Wake Forest University (P30AG021332) and the Clinical and Translational Science Institutes, funded by the National Center for Advancing Translational Science at Wake Forest University (UL10TR001420).

## Authors’ contributions

Peggy M Cawthon, Paul M Coen, Steven R Cummings, Steven B Kritchevsky, Anne B Newman, Russell T Hepple, Karyn Esser and Gregory J Tranah designed the study. Paul M Coen and Gregory J Tranah drafted the manuscript. Haley N Barnes, Paul M Coen, Daniel S Evans, and Zhiguang Huo, conducted the genetic analyses. Paul M Coen, Steven R Cummings, Steven B Kritchevsky, Anne B Newman, Russell T Hepple, Karyn Esser, Olaya Santiago Fernandez and Ana Maria Cuervo and Gregory J Tranah contributed to the interpretation of the results. Peggy M Cawthon, Paul M Coen, Frederico GS Toledo, Steven B Kritchevsky, Anne B Newman, Steven R Cummings, Gregory J Tranah have critically revised the manuscript. All authors reviewed and approved the final version of the manuscript and Haley N Barnes and Zhiguang Huo had full access to the data in the study and accept responsibility to submit for publication.

## Data availability statement

All SOMMA data are publicly available via a web portal. Updated datasets are released approximately every 6 months (https://www.sommastudy.com/for-investigators).

## SUPPLEMENTARY INFORMATION

**Supplementary Table 1:** RNAseq quality metrics for each sample, including: number of raw pairs, alignment rate, number of aligned reads, duplication rate, and number of aligned reads deduplicated.

| Sample | Number of raw pairs | Alignment Rate | Number of aligned reads | Duplication Rate | Number of aligned reads deduplicated |
| --- | --- | --- | --- | --- | --- |
| 1 | 96393453 | 95.67% | 184441951 | 78.70% | 39359245 |
| 2 | 100059418 | 96.13% | 192367958 | 68.10% | 61433169 |
| 3 | 93532566 | 96.08% | 179727375 | 68.50% | 56555649 |
| 4 | 90130020 | 95.93% | 172917121 | 62.90% | 64215917 |
| 5 | 93513176 | 95.74% | 179061926 | 66.50% | 60023883 |
| 6 | 93399507 | 96.03% | 179390873 | 78.60% | 38357663 |
| 7 | 91533891 | 96.04% | 175814274 | 75.50% | 43079694 |
| 8 | 95660352 | 95.15% | 182046440 | 86% | 25426123 |
| 9 | 92869196 | 95.60% | 177559973 | 73.80% | 46546318 |
| 10 | 94206508 | 95.61% | 180140177 | 65.40% | 62368154 |
| 11 | 91580510 | 96.24% | 176269851 | 68.30% | 55963786 |
| 12 | 96476311 | 96.31% | 185835269 | 65.40% | 64275793 |
| 13 | 93256753 | 96.06% | 179170646 | 69.90% | 53926180 |
| 14 | 94737694 | 96.51% | 182871302 | 70.80% | 53335997 |
| 15 | 94853873 | 94.61% | 179484404 | 91.60% | 15094298 |
| 16 | 93254346 | 96.10% | 179236781 | 79.80% | 36214863 |
| 17 | 94768451 | 96.06% | 182078220 | 70.30% | 53994168 |
| 18 | 92703785 | 96.36% | 178650402 | 61.20% | 69327789 |
| 19 | 92792742 | 96.33% | 178781101 | 65.80% | 61190811 |
| 20 | 95448973 | 96.50% | 184214920 | 71.60% | 52242426 |
| 21 | 96386549 | 95.70% | 184490763 | 65.20% | 64208390 |
| 22 | 94049712 | 95.77% | 180144045 | 85.40% | 26374250 |
| 23 | 92534259 | 95.52% | 176781852 | 82% | 31751068 |
| 24 | 96136369 | 95.47% | 183567705 | 77.70% | 40894545 |
| 25 | 92684795 | 96.66% | 179169760 | 79% | 37553138 |
| 26 | 94274385 | 95.62% | 180291886 | 81% | 34329087 |
| 27 | 95238852 | 95.32% | 181565967 | 81.80% | 32954237 |
| 28 | 94658476 | 95.26% | 180343006 | 89.90% | 18235077 |
| 29 | 92917699 | 95.43% | 177341522 | 75.20% | 43975639 |
| 30 | 97826542 | 96.21% | 188243115 | 72.50% | 51785757 |
| 31 | 93211235 | 95.45% | 177942884 | 83.40% | 29533846 |
| 32 | 95606463 | 95.69% | 182973610 | 75.20% | 45433035 |
| 33 | 95441961 | 96.31% | 183849801 | 74.60% | 46701458 |
| 34 | 96075319 | 96.12% | 184697195 | 74.20% | 47673981 |
| 35 | 95074060 | 96.35% | 183208134 | 70.80% | 53417857 |
| 36 | 91693188 | 95.82% | 175727627 | 77.60% | 39393394 |
| 37 | 92617017 | 92.67% | 171647448 | 90.10% | 16974404 |
| 38 | 95760744 | 95.74% | 183363658 | 78% | 40363306 |
| 39 | 93959617 | 94.95% | 178419945 | 82.10% | 32008520 |
| 40 | 97687230 | 95.69% | 186945742 | 72.80% | 50891281 |
| 41 | 95290985 | 96.08% | 183118608 | 76.10% | 43755133 |
| 42 | 96405400 | 96.10% | 185288591 | 71.30% | 53153408 |
| 43 | 91618248 | 95.70% | 175361499 | 74% | 45638953 |
| 44 | 90604635 | 95.59% | 173221766 | 79.70% | 35235993 |
| 45 | 91908525 | 94.24% | 173221121 | 73.90% | 45147052 |
| 46 | 93339668 | 95.45% | 178183219 | 86.60% | 23847280 |
| 47 | 93228195 | 95.38% | 177838787 | 80.10% | 35379447 |
| 48 | 94185936 | 95.70% | 180277596 | 85.90% | 25478974 |
| 49 | 93076777 | 95.60% | 177965955 | 74.90% | 44623024 |
| 50 | 92853231 | 95.51% | 177372625 | 67.50% | 57638727 |
| 51 | 95924802 | 95.83% | 183844971 | 83.20% | 30890475 |
| 52 | 91701902 | 93.61% | 171676642 | 88.50% | 19794352 |
| 53 | 93978044 | 95.60% | 179680494 | 82.20% | 31952553 |
| 54 | 93498088 | 94.84% | 177339169 | 85.90% | 25052790 |
| 55 | 91373701 | 95.10% | 173792287 | 86% | 24339052 |
| 56 | 92633161 | 94.21% | 174537705 | 92.40% | 13282764 |
| 57 | 93632029 | 94.82% | 177555337 | 74.70% | 44895907 |
| 58 | 96935810 | 96.11% | 186322556 | 66.40% | 62655363 |
| 59 | 95970326 | 95.97% | 184214134 | 68.70% | 57608266 |
| 60 | 92800167 | 95.46% | 177179393 | 86.60% | 23666468 |
| 61 | 102699034 | 96.30% | 197801041 | 64.90% | 69504546 |
| 62 | 92654946 | 94.91% | 175870882 | 85% | 26296527 |
| 63 | 92637261 | 94.25% | 174620099 | 85.40% | 25547736 |
| 64 | 93933534 | 94.91% | 178301144 | 86% | 24893302 |
| 65 | 94850969 | 94.69% | 179622664 | 91.10% | 15967433 |
| 66 | 93576231 | 95.41% | 178562087 | 80.90% | 34065450 |
| 67 | 90485671 | 95.26% | 172384541 | 75.90% | 41587116 |
| 68 | 93379297 | 94.19% | 175909909 | 91.20% | 15397217 |
| 69 | 96498988 | 95.91% | 185101771 | 73.10% | 49753846 |
| 70 | 71461019 | 95.40% | 136344256 | 59.30% | 55504737 |
| 71 | 93752448 | 95.60% | 179248936 | 59.20% | 73073568 |
| 72 | 86411126 | 96.23% | 166306753 | 58.30% | 69376925 |
| 73 | 83235259 | 96.26% | 160250568 | 55.50% | 71262583 |
| 74 | 81261596 | 96.36% | 156615091 | 60.10% | 62471112 |
| 75 | 97783727 | 95.85% | 187450816 | 57.10% | 80348879 |
| 76 | 83767720 | 95.75% | 160407526 | 57.60% | 68092398 |
| 77 | 86729806 | 95.31% | 165318863 | 57.60% | 70036456 |
| 78 | 110942864 | 95.51% | 211919823 | 57% | 91162871 |
| 79 | 104966827 | 96.19% | 201945250 | 56% | 88772846 |
| 80 | 98253075 | 96.48% | 189598150 | 65.60% | 65198970 |
| 81 | 116927639 | 96.27% | 225129926 | 56.50% | 97959452 |
| 82 | 112176949 | 96.19% | 215797160 | 58.40% | 89813138 |
| 83 | 98534039 | 95.80% | 188796585 | 53.50% | 87847469 |
| 84 | 108954720 | 95.86% | 208892196 | 53.40% | 97416327 |
| 85 | 84097255 | 95.78% | 161090321 | 57.10% | 69129756 |
| 86 | 95236027 | 95.90% | 182663085 | 61.60% | 70206810 |
| 87 | 88597819 | 95.87% | 169884224 | 55.30% | 75955704 |
| 88 | 96328455 | 96.12% | 185180033 | 57.80% | 78177200 |
| 89 | 90461109 | 96.09% | 173855662 | 56.80% | 75088574 |
| 90 | 102870253 | 95.60% | 196695477 | 57.10% | 84377364 |
| 91 | 108655599 | 96.07% | 208762677 | 55% | 93925156 |
| 92 | 106334623 | 95.39% | 202873100 | 58% | 85169516 |
| 93 | 95273543 | 96.52% | 183917484 | 59.50% | 74534598 |
| 94 | 92910303 | 95.10% | 176722146 | 61.70% | 67734224 |
| 95 | 85877520 | 95.74% | 164431755 | 57.30% | 70181106 |
| 96 | 101420521 | 96.01% | 194746668 | 57.80% | 82270980 |
| 97 | 106560498 | 95.79% | 204151071 | 60.10% | 81465013 |
| 98 | 88052217 | 95.85% | 168790927 | 61.40% | 65182021 |
| 99 | 86639281 | 95.98% | 166305148 | 60.20% | 66160365 |
| 100 | 110166267 | 96.12% | 211781602 | 58.40% | 88101366 |
| 101 | 68517291 | 95.90% | 131415364 | 58.40% | 54667509 |
| 102 | 82185872 | 89.49% | 147099825 | 56.90% | 63463056 |
| 103 | 81881951 | 91.49% | 149826924 | 51.60% | 72577249 |
| 104 | 79982310 | 92.80% | 148454721 | 54.20% | 68030223 |
| 105 | 80404905 | 93.43% | 150237431 | 54.10% | 68971717 |
| 106 | 80821283 | 92.89% | 150151720 | 56.80% | 64805299 |
| 107 | 81203942 | 92.14% | 149636180 | 52.50% | 71064306 |
| 108 | 83124572 | 93.65% | 155692925 | 55% | 70035174 |
| 109 | 83732758 | 91.81% | 153750133 | 56% | 67697013 |
| 110 | 85425657 | 93.21% | 159252490 | 55.20% | 71368045 |
| 111 | 83608132 | 92.59% | 154821249 | 55.10% | 69477695 |
| 112 | 80057039 | 92.61% | 148280864 | 51.40% | 72014599 |
| 113 | 80421460 | 92.78% | 149231205 | 55.70% | 66145867 |
| 114 | 79661371 | 93.39% | 148792790 | 52.40% | 70837393 |
| 115 | 78552545 | 93.67% | 147161472 | 49.60% | 74116342 |
| 116 | 80742166 | 93.70% | 151310619 | 53.80% | 69932072 |
| 117 | 81952954 | 93.45% | 153170108 | 51.40% | 74467136 |
| 118 | 84605721 | 92.48% | 156488222 | 53.50% | 72754366 |
| 119 | 83317876 | 93.43% | 155693857 | 54.60% | 70715421 |
| 120 | 84072070 | 92.31% | 155213840 | 55.10% | 69658730 |
| 121 | 78879445 | 93.85% | 148060431 | 54.30% | 67613149 |
| 122 | 81065245 | 93.13% | 150987465 | 52.90% | 71065006 |
| 123 | 80217494 | 93.74% | 150386891 | 52.60% | 71283062 |
| 124 | 78825058 | 93.19% | 146907604 | 53% | 69007099 |
| 125 | 82173370 | 92.22% | 151561681 | 56% | 66722801 |
| 126 | 84325245 | 91.89% | 154967259 | 55.80% | 68529272 |
| 127 | 83005575 | 92.12% | 152925352 | 52.40% | 72862136 |
| 128 | 79764300 | 92.28% | 147216468 | 54.60% | 66881432 |
| 129 | 80033284 | 93.50% | 149658311 | 53.60% | 69425984 |
| 130 | 83094845 | 92.07% | 153013625 | 56% | 67339202 |
| 131 | 81833515 | 93.24% | 152604746 | 53.20% | 71429012 |
| 132 | 81767329 | 92.75% | 151672136 | 54.50% | 69022261 |
| 133 | 86613650 | 93.83% | 162536626 | 50.90% | 79868855 |
| 134 | 84575964 | 93.04% | 157374691 | 52.80% | 74277949 |
| 135 | 84551330 | 93.20% | 157608706 | 57.30% | 67364855 |
| 136 | 79377942 | 92.20% | 146379254 | 54.60% | 66496956 |
| 137 | 82220781 | 93.55% | 153841128 | 53.80% | 71087389 |
| 138 | 84490367 | 93.66% | 158262891 | 49% | 80737589 |
| 139 | 79135730 | 93.46% | 147925972 | 54.60% | 67200337 |
| 140 | 79315800 | 93.29% | 147985482 | 53.80% | 68321650 |
| 141 | 83183251 | 93.78% | 156011569 | 56.20% | 68318618 |
| 142 | 80033861 | 93.20% | 149184350 | 54.50% | 67886689 |
| 143 | 81365287 | 93.54% | 152211890 | 53.10% | 71334270 |
| 144 | 78302039 | 93.65% | 146653385 | 52% | 70451521 |
| 145 | 78250641 | 93.72% | 146677528 | 50.20% | 72978169 |
| 146 | 88982141 | 93.56% | 166509772 | 51.30% | 81144529 |
| 147 | 83465583 | 93.76% | 156512508 | 52.70% | 73991949 |
| 148 | 78062842 | 93.16% | 145453918 | 54.90% | 65605230 |
| 149 | 82988051 | 93.61% | 155366184 | 54.10% | 71328165 |
| 150 | 81862895 | 93.18% | 152566757 | 53.50% | 70874912 |
| 151 | 83357795 | 92.99% | 155033190 | 53.90% | 71547631 |
| 152 | 78541474 | 92.61% | 145477010 | 55.20% | 65238613 |
| 153 | 79658492 | 93.21% | 148502581 | 54% | 68314118 |
| 154 | 83568623 | 93.12% | 155630779 | 51.10% | 76070555 |
| 155 | 82216893 | 93.36% | 153507981 | 54.80% | 69417332 |
| 156 | 80284841 | 93.01% | 149342427 | 53.80% | 69032431 |
| 157 | 81855521 | 92.63% | 151649262 | 57.70% | 64177810 |
| 158 | 83777511 | 92.15% | 154404604 | 56.40% | 67384639 |
| 159 | 76397124 | 92.84% | 141852133 | 52.10% | 67974888 |
| 160 | 76699266 | 93.64% | 143635884 | 52.50% | 68190049 |
| 161 | 81119697 | 92.98% | 150848135 | 57.30% | 64381281 |
| 162 | 80828343 | 93.88% | 151762556 | 53.70% | 70201572 |
| 163 | 81754170 | 92.54% | 151315363 | 54.40% | 68934509 |
| 164 | 77182270 | 92.88% | 143378696 | 54.50% | 65188366 |
| 165 | 83277951 | 91.93% | 153115251 | 54.80% | 69242593 |
| 166 | 81434647 | 93.24% | 151858802 | 53% | 71313059 |
| 167 | 80097377 | 93.07% | 149095520 | 56% | 65587909 |
| 168 | 80351222 | 92.75% | 149049068 | 55.40% | 66477867 |
| 169 | 81065470 | 92.60% | 150136734 | 56.50% | 65376792 |
| 170 | 79732292 | 92.77% | 147932802 | 51.20% | 72190979 |
| 171 | 81364923 | 92.82% | 151041094 | 54.70% | 68468523 |
| 172 | 84217555 | 92.71% | 156156243 | 53.40% | 72719070 |
| 173 | 82659700 | 91.86% | 151855951 | 58.90% | 62427158 |
| 174 | 77764144 | 92.67% | 144122794 | 54.70% | 65326768 |
| 175 | 78458276 | 93.61% | 146894807 | 53.40% | 68434532 |
| 176 | 82281245 | 92.59% | 152375583 | 54.70% | 69096970 |
| 177 | 81723750 | 93.04% | 152071478 | 53.70% | 70467804 |
| 178 | 92966002 | 93.08% | 173061954 | 53% | 81409313 |
| 179 | 108642167 | 92.90% | 201857878 | 54.50% | 91931884 |
| 180 | 110042963 | 93.11% | 204915230 | 55.40% | 91326091 |
| 181 | 80782419 | 91.74% | 148214005 | 55.40% | 66133720 |
| 182 | 84525342 | 93.26% | 157656520 | 54.20% | 72184934 |
| 183 | 83707351 | 92.93% | 155580510 | 56.70% | 67374897 |
| 184 | 81396801 | 92.67% | 150863483 | 54.20% | 69072120 |
| 185 | 79461855 | 92.93% | 147683370 | 53.60% | 68516893 |
| 186 | 82835786 | 93.45% | 154824888 | 56.10% | 68011860 |
| 187 | 80121046 | 91.52% | 146653254 | 56.20% | 64198018 |
| 188 | 80677720 | 93.08% | 150191167 | 51.70% | 72467988 |
| 189 | 73976840 | 92.95% | 137522860 | 55.50% | 61139754 |
| 190 | 84296590 | 92.68% | 156244470 | 54.30% | 71397717 |
| 191 | 79357053 | 92.05% | 146094379 | 57.40% | 62260719 |
| 192 | 83738743 | 92.78% | 155381196 | 55.10% | 69733453 |
| 193 | 81718359 | 93.00% | 151991385 | 55.70% | 67276304 |
| 194 | 82041039 | 92.73% | 152145911 | 53.70% | 70411580 |
| 195 | 80029302 | 93.27% | 149286808 | 54.60% | 67736131 |
| 196 | 80785074 | 92.28% | 149102807 | 58.20% | 62289713 |
| 197 | 83843652 | 89.95% | 150834458 | 55.60% | 66959638 |
| 198 | 83590981 | 90.54% | 151374766 | 54.40% | 69016209 |
| 199 | 83437723 | 90.56% | 151119766 | 61.30% | 58460828 |
| 200 | 83200552 | 90.82% | 151130394 | 58.30% | 63046406 |
| 201 | 85626471 | 91.26% | 156284569 | 54.70% | 70851462 |
| 202 | 85639782 | 91.15% | 156127532 | 55.30% | 69759580 |
| 203 | 82537377 | 92.16% | 152128868 | 54.80% | 68808968 |
| 204 | 85570850 | 91.07% | 155854052 | 55.50% | 69431392 |
| 205 | 85934669 | 88.37% | 151874286 | 57.70% | 64291224 |
| 206 | 82777122 | 89.44% | 148070369 | 56.30% | 64655527 |
| 207 | 84965709 | 91.77% | 155946997 | 60.90% | 60917009 |
| 208 | 83933360 | 91.66% | 153860969 | 56.30% | 67229061 |
| 209 | 84894553 | 91.66% | 155625165 | 54.60% | 70729417 |
| 210 | 86259859 | 91.23% | 157396509 | 57.40% | 67053274 |
| 211 | 84346300 | 90.21% | 152170674 | 60.30% | 60352643 |
| 212 | 81677653 | 92.44% | 151003784 | 54% | 69511530 |
| 213 | 84048781 | 90.99% | 152954587 | 54.50% | 69606002 |
| 214 | 83654135 | 91.72% | 153456971 | 56.10% | 67299283 |
| 215 | 82130420 | 91.59% | 150453661 | 55.80% | 66435733 |
| 216 | 86286063 | 92.62% | 159833996 | 55.20% | 71548012 |
| 217 | 81626143 | 92.37% | 150800166 | 53.50% | 70150436 |
| 218 | 88346127 | 92.33% | 163143773 | 61.10% | 63421969 |
| 219 | 84865918 | 91.48% | 155278413 | 59.90% | 62191416 |
| 220 | 84973197 | 91.01% | 154664003 | 57.10% | 66420352 |
| 221 | 85254795 | 91.64% | 156259490 | 52.70% | 73942216 |
| 222 | 83789654 | 91.01% | 152511134 | 58.80% | 62864169 |
| 223 | 95815599 | 92.47% | 177201844 | 57.80% | 74694621 |
| 224 | 81589169 | 92.29% | 150602813 | 53.40% | 70253955 |
| 225 | 84008407 | 93.45% | 157012490 | 54.40% | 71645670 |
| 226 | 91458190 | 92.84% | 169827777 | 52% | 81521417 |
| 227 | 85816384 | 92.18% | 158208155 | 51.90% | 76103627 |
| 228 | 85124387 | 92.51% | 157502606 | 53.30% | 73491302 |
| 229 | 95718199 | 92.66% | 177393846 | 50.90% | 87012401 |
| 230 | 88765558 | 91.84% | 163040208 | 50.20% | 81131345 |
| 231 | 85307518 | 92.10% | 157139563 | 47.20% | 82950928 |
| 232 | 85215525 | 92.17% | 157078878 | 52.20% | 75159281 |
| 233 | 92867670 | 92.78% | 172320565 | 50.30% | 85711818 |
| 234 | 95802997 | 92.10% | 176464214 | 51% | 86392608 |
| 235 | 90056750 | 92.42% | 166462816 | 52.40% | 79208945 |
| 236 | 88674254 | 92.26% | 163617746 | 49.10% | 83319295 |
| 237 | 87963713 | 93.34% | 164213747 | 49.40% | 83028930 |
| 238 | 92809866 | 92.70% | 172061848 | 51.70% | 83186393 |
| 239 | 87668758 | 92.36% | 161935944 | 54.80% | 73257571 |
| 240 | 83691132 | 93.13% | 155876722 | 52.30% | 74285916 |
| 241 | 87620635 | 93.43% | 163722214 | 48.50% | 84303328 |
| 242 | 87370476 | 93.83% | 163955893 | 51.50% | 79457633 |
| 243 | 85183296 | 93.51% | 159304394 | 48.90% | 81388632 |
| 244 | 85305143 | 91.00% | 155259108 | 59.40% | 62964001 |
| 245 | 87686269 | 91.76% | 160913835 | 50.10% | 80229213 |
| 246 | 85759147 | 92.88% | 159301455 | 45.80% | 86380616 |
| 247 | 86761827 | 92.46% | 160444212 | 52.30% | 76597750 |
| 248 | 95574789 | 93.14% | 178043997 | 53.60% | 82642295 |
| 249 | 84799240 | 91.03% | 154390548 | 58.90% | 63417450 |
| 250 | 92994203 | 92.25% | 171580708 | 54.90% | 77303002 |
| 251 | 90002275 | 93.51% | 168319214 | 52.50% | 79979804 |
| 252 | 93496332 | 92.41% | 172791290 | 53.20% | 80942510 |
| 253 | 93429096 | 93.13% | 174026090 | 53.40% | 81155282 |
| 254 | 92210444 | 93.87% | 173118774 | 49.60% | 87229532 |
| 255 | 86830309 | 93.01% | 161524479 | 51.20% | 78790992 |
| 256 | 85668489 | 93.69% | 160529297 | 53.20% | 75101647 |
| 257 | 82811822 | 93.50% | 154851807 | 56.70% | 67098167 |
| 258 | 92210607 | 92.75% | 171043151 | 56.20% | 74920206 |
| 259 | 86670973 | 92.98% | 161175596 | 54% | 74204742 |
| 260 | 85317863 | 92.59% | 157985436 | 53.60% | 73349237 |
| 261 | 84376356 | 93.29% | 157430753 | 52% | 75551090 |
| 262 | 88846406 | 93.69% | 166474759 | 52.80% | 78535808 |
| 263 | 84250864 | 93.37% | 157323970 | 49.90% | 78810698 |
| 264 | 83884503 | 93.41% | 156707758 | 55.70% | 69362822 |
| 265 | 84039086 | 93.25% | 156727952 | 58.10% | 65613249 |
| 266 | 85755986 | 93.72% | 160745223 | 52.40% | 76444502 |
| 267 | 84323917 | 93.02% | 156873725 | 55.10% | 70465299 |
| 268 | 86565553 | 93.53% | 161926567 | 50.80% | 79604802 |
| 269 | 86567025 | 93.16% | 161289477 | 50.10% | 80523325 |
| 270 | 88494339 | 92.87% | 164361568 | 52% | 78929966 |
| 271 | 86623009 | 93.02% | 161151152 | 51.20% | 78600588 |
| 272 | 86327648 | 93.10% | 160745980 | 54% | 73918147 |
| 273 | 83141346 | 93.24% | 155036034 | 53.30% | 72386235 |
| 274 | 83302444 | 93.24% | 155343331 | 60.30% | 61680278 |
| 275 | 84909067 | 93.47% | 158736296 | 53.10% | 74517144 |
| 276 | 94998934 | 93.04% | 176782669 | 54.20% | 80973713 |
| 277 | 85473106 | 91.72% | 156783969 | 48% | 81529587 |
| 278 | 84419283 | 93.08% | 157151408 | 49.60% | 79147797 |
| 279 | 84339155 | 92.54% | 156093345 | 54.10% | 71659275 |
| 280 | 82898656 | 93.49% | 155005772 | 57.10% | 66469529 |
| 281 | 88618338 | 92.36% | 163687680 | 55.50% | 72864712 |
| 282 | 84028068 | 92.83% | 155998753 | 55.40% | 69596791 |
| 283 | 86880065 | 92.54% | 160795831 | 54.20% | 73623117 |
| 284 | 84845899 | 93.53% | 158711551 | 51.20% | 77485314 |
| 285 | 84860629 | 92.11% | 156330197 | 52.40% | 74379075 |
| 286 | 84257108 | 93.42% | 157421881 | 52.40% | 74920353 |
| 287 | 84280613 | 93.66% | 157878588 | 50.10% | 78725756 |
| 288 | 84861735 | 93.52% | 158721939 | 53.70% | 73496627 |
| 289 | 85386584 | 93.20% | 159158664 | 51.50% | 77183783 |
| 290 | 85547278 | 91.97% | 157357004 | 64.40% | 56086510 |
| 291 | 85506856 | 92.17% | 157623630 | 50.60% | 77883014 |
| 292 | 87465435 | 92.87% | 162465151 | 48.70% | 83288893 |
| 293 | 89476865 | 93.44% | 167220449 | 50.30% | 83141110 |
| 294 | 82110594 | 93.53% | 153591414 | 55% | 69078281 |
| 295 | 84093778 | 92.59% | 155729679 | 60.40% | 61678538 |
| 296 | 83423489 | 93.01% | 155185118 | 59.60% | 62749001 |
| 297 | 85932540 | 93.29% | 160337553 | 59.40% | 65131305 |
| 298 | 83538416 | 93.34% | 155953809 | 52.40% | 74289150 |
| 299 | 84091188 | 93.52% | 157277665 | 49.20% | 79941873 |
| 300 | 90069946 | 93.01% | 167540972 | 52.10% | 80254592 |
| 301 | 84061111 | 93.66% | 157457891 | 50.80% | 77394403 |
| 302 | 83155272 | 93.16% | 154932286 | 57.50% | 65789921 |
| 303 | 83585064 | 92.55% | 154711276 | 56.30% | 67639225 |
| 304 | 84628574 | 93.46% | 158188844 | 55.90% | 69742868 |
| 305 | 83011782 | 92.14% | 152977493 | 54.20% | 70059215 |
| 306 | 81280441 | 93.78% | 152451484 | 50.80% | 74998130 |
| 307 | 84245261 | 92.76% | 156299058 | 54% | 71892451 |
| 308 | 84725802 | 92.59% | 156893654 | 51.20% | 76591677 |
| 309 | 84866329 | 92.66% | 157282114 | 52.60% | 74575212 |
| 310 | 85688224 | 92.84% | 159099875 | 53.30% | 74358069 |
| 311 | 85437828 | 91.87% | 156986962 | 62.70% | 58478149 |
| 312 | 84433153 | 92.86% | 156816220 | 55.70% | 69523283 |
| 313 | 84621049 | 92.92% | 157251935 | 53.20% | 73551058 |
| 314 | 83144014 | 93.91% | 156162315 | 50.30% | 77646238 |
| 315 | 83362769 | 92.86% | 154822328 | 50.70% | 76272766 |
| 316 | 84481479 | 93.31% | 157655895 | 49.30% | 79968258 |
| 317 | 86230336 | 93.73% | 161655502 | 50.80% | 79536622 |
| 318 | 84799431 | 92.73% | 157261019 | 49.80% | 78957526 |
| 319 | 78405835 | 93.18% | 146109354 | 48% | 75928263 |
| 320 | 88892787 | 93.76% | 166692480 | 50.80% | 82020430 |
| 321 | 85070862 | 93.32% | 158769321 | 48% | 82521937 |
| 322 | 84947408 | 94.30% | 160214849 | 50.80% | 78829492 |
| 323 | 92407695 | 93.73% | 173228551 | 52.50% | 82368708 |
| 324 | 84891874 | 94.27% | 160048590 | 50.70% | 78926705 |
| 325 | 85709910 | 94.39% | 161804370 | 51.50% | 78417315 |
| 326 | 83101247 | 94.01% | 156241366 | 48.50% | 80410232 |
| 327 | 82308793 | 93.72% | 154274395 | 52.10% | 73843575 |
| 328 | 87109225 | 94.00% | 163759092 | 51% | 80301894 |
| 329 | 86532923 | 92.20% | 159572561 | 50.90% | 78415267 |
| 330 | 90737882 | 90.54% | 164315402 | 54.90% | 74053890 |
| 331 | 83355319 | 92.29% | 153862208 | 50.30% | 76523033 |
| 332 | 85026830 | 93.60% | 159169119 | 51.60% | 76985647 |
| 333 | 83133743 | 92.89% | 154448461 | 52.50% | 73314153 |
| 334 | 81974796 | 92.52% | 151679531 | 49.70% | 76328624 |
| 335 | 85411349 | 93.16% | 159131845 | 48.70% | 81630732 |
| 336 | 87230169 | 89.93% | 156896035 | 51.20% | 76597809 |
| 337 | 86522843 | 93.28% | 161421146 | 52.10% | 77300193 |
| 338 | 83912636 | 92.91% | 155918473 | 47.40% | 81939343 |
| 339 | 96064117 | 93.21% | 179087734 | 55.50% | 79780353 |
| 340 | 91874480 | 92.19% | 169398960 | 54.10% | 77734137 |
| 341 | 90458184 | 93.45% | 169069924 | 54.20% | 77368105 |
| 342 | 92262874 | 92.32% | 170355177 | 56.30% | 74361329 |
| 343 | 88623713 | 93.33% | 165428691 | 54.40% | 75387321 |
| 344 | 87207805 | 91.69% | 159918728 | 62.10% | 60610291 |
| 345 | 91004676 | 91.07% | 165761485 | 65.50% | 57159741 |
| 346 | 90916275 | 93.80% | 170560323 | 54.10% | 78288436 |
| 347 | 82776921 | 91.55% | 151571920 | 65.70% | 52028736 |
| 348 | 92768155 | 91.99% | 170681073 | 52.80% | 80587319 |
| 349 | 87856604 | 91.93% | 161536091 | 56% | 71121274 |
| 350 | 94246257 | 93.71% | 176642932 | 54% | 81343271 |
| 351 | 90609020 | 92.28% | 167228668 | 60.30% | 66404884 |
| 352 | 96498298 | 92.76% | 179016244 | 60% | 71599729 |
| 353 | 90315456 | 92.62% | 167293252 | 57.70% | 70834030 |
| 354 | 91413142 | 91.00% | 166373604 | 59.60% | 67260982 |
| 355 | 87915585 | 92.42% | 162503027 | 61.70% | 62305118 |
| 356 | 91125683 | 92.91% | 169336075 | 57.40% | 72191861 |
| 357 | 91398473 | 92.76% | 169571520 | 54.20% | 77661393 |
| 358 | 95481994 | 91.81% | 175329119 | 60.40% | 69382289 |
| 359 | 110818214 | 92.60% | 205241457 | 57.60% | 87117111 |
| 360 | 94806068 | 92.95% | 176246636 | 54.70% | 79789029 |
| 361 | 89836893 | 92.71% | 166580194 | 56.10% | 73142972 |
| 362 | 98638711 | 93.23% | 183921845 | 56.70% | 79659069 |
| 363 | 93853377 | 92.12% | 172917042 | 55.50% | 77019448 |
| 364 | 100275484 | 91.79% | 184078267 | 58.40% | 76604986 |
| 365 | 92495429 | 93.01% | 172062371 | 54.80% | 77718704 |
| 366 | 88715899 | 92.84% | 164731494 | 55.50% | 73260860 |
| 367 | 92461693 | 93.31% | 172556715 | 55.80% | 76329898 |
| 368 | 102100512 | 92.74% | 189382528 | 54% | 87061978 |
| 369 | 90504044 | 92.70% | 167798855 | 60.10% | 66899554 |
| 370 | 89709289 | 93.94% | 168552896 | 52.20% | 80537509 |
| 371 | 87126691 | 93.41% | 162764208 | 60.10% | 64993578 |
| 372 | 84451594 | 93.81% | 158447618 | 54.30% | 72379951 |
| 373 | 92484999 | 93.30% | 172582788 | 56.20% | 75514636 |
| 374 | 95069991 | 92.37% | 175627025 | 53.60% | 81522149 |
| 375 | 91417795 | 93.66% | 171251489 | 54.30% | 78191020 |
| 376 | 87960991 | 90.90% | 159916106 | 59.70% | 64483465 |
| 377 | 99886475 | 93.44% | 186660828 | 57.40% | 79514036 |
| 378 | 87890978 | 92.47% | 162553511 | 57.30% | 69367802 |
| 379 | 92017579 | 93.07% | 171275765 | 61.20% | 66437963 |
| 380 | 87881414 | 93.31% | 163996843 | 54.10% | 75296785 |
| 381 | 88237604 | 93.83% | 165581376 | 52.10% | 79392676 |
| 382 | 87808127 | 93.01% | 163340130 | 55.70% | 72401527 |
| 383 | 91892021 | 94.02% | 172792672 | 52.20% | 82574586 |
| 384 | 86824993 | 93.68% | 162683119 | 53.70% | 75385841 |
| 385 | 83169708 | 92.44% | 153768393 | 64.50% | 54605416 |
| 386 | 92957654 | 91.89% | 170829216 | 61.60% | 65562022 |
| 387 | 90627760 | 93.92% | 170231373 | 63.60% | 61998824 |
| 388 | 94159915 | 93.71% | 176478389 | 53.60% | 81955000 |
| 389 | 93895734 | 92.31% | 173357587 | 67% | 57291911 |
| 390 | 93110943 | 91.98% | 171288798 | 65.50% | 59067248 |
| 391 | 91179562 | 93.39% | 170307295 | 60.70% | 66994666 |
| 392 | 94007538 | 94.13% | 176984606 | 57% | 76037137 |
| 393 | 92783466 | 91.90% | 170532129 | 65.30% | 59175784 |
| 394 | 93347423 | 93.30% | 174193583 | 54.10% | 80031876 |
| 395 | 93719900 | 92.07% | 172581611 | 56.80% | 74499816 |
| 396 | 94289838 | 91.92% | 173338731 | 68.30% | 54872025 |
| 397 | 94653775 | 93.44% | 176883243 | 62.80% | 65720019 |
| 398 | 94164370 | 93.32% | 175752605 | 61.90% | 66968050 |
| 399 | 88567436 | 93.30% | 165258210 | 69.10% | 51033967 |
| 400 | 98416526 | 92.21% | 181506660 | 61% | 70857795 |
| 401 | 95476259 | 92.08% | 175822231 | 74% | 45652965 |
| 402 | 92721577 | 93.59% | 173558028 | 66.90% | 57505064 |
| 403 | 92259150 | 91.10% | 168094982 | 66.80% | 55805579 |
| 404 | 88083362 | 93.76% | 165177022 | 58.10% | 69219073 |
| 405 | 92673640 | 92.23% | 170944821 | 68.20% | 54331377 |
| 406 | 91470878 | 90.71% | 165950945 | 68.30% | 52630329 |
| 407 | 92289829 | 91.24% | 168411040 | 66.50% | 56472297 |
| 408 | 88452199 | 92.23% | 163152205 | 63.10% | 60181547 |
| 409 | 94457069 | 91.78% | 173385603 | 69.20% | 53418541 |
| 410 | 90804922 | 90.60% | 164542909 | 59.40% | 66759518 |
| 411 | 92170656 | 92.31% | 170159210 | 58.80% | 70080399 |
| 412 | 88015234 | 90.63% | 159541716 | 67.40% | 52066946 |
| 413 | 90896266 | 92.02% | 167291419 | 66.50% | 55964132 |
| 414 | 95267470 | 93.90% | 178918725 | 63.10% | 66019900 |
| 415 | 90307683 | 89.91% | 162395740 | 75.20% | 40311703 |
| 416 | 89954030 | 93.51% | 168237445 | 72.70% | 45963463 |
| 417 | 92393129 | 90.77% | 167722925 | 58.60% | 69448174 |
| 418 | 96504239 | 92.38% | 178305273 | 54.50% | 81161684 |
| 419 | 94181054 | 92.24% | 173736160 | 53.60% | 80633299 |
| 420 | 88132332 | 92.81% | 163596957 | 55.20% | 73288089 |
| 421 | 92289521 | 93.42% | 172434278 | 56.30% | 75417880 |
| 422 | 93722558 | 91.93% | 172308930 | 59.10% | 70468747 |
| 423 | 91533009 | 90.67% | 165985429 | 72.30% | 45947853 |
| 424 | 98072046 | 92.20% | 180847605 | 66.80% | 59997635 |
| 425 | 94242577 | 88.97% | 167693401 | 61.30% | 64908370 |
| 426 | 90280373 | 91.21% | 164689198 | 66.60% | 54965722 |
| 427 | 93086902 | 94.13% | 175245153 | 62.80% | 65237940 |
| 428 | 88128911 | 92.82% | 163601591 | 63.40% | 59947062 |
| 429 | 93573424 | 93.78% | 175506311 | 62.50% | 65822250 |
| 430 | 93868423 | 93.42% | 175381316 | 57.60% | 74397340 |
| 431 | 98482895 | 90.88% | 179010692 | 73.80% | 46966176 |
| 432 | 93638980 | 93.02% | 174207479 | 64.10% | 62462076 |
| 433 | 89719416 | 93.27% | 167369730 | 65.70% | 57463722 |
| 434 | 95073085 | 91.95% | 174833170 | 61.70% | 66880236 |
| 435 | 89822530 | 92.97% | 167011901 | 67.30% | 54612076 |
| 436 | 91834032 | 92.67% | 170198511 | 61.20% | 66085430 |
| 437 | 92515385 | 89.52% | 165636384 | 71.80% | 46708002 |
| 438 | 81638563 | 93.65% | 152909707 | 55.20% | 68573636 |
| 439 | 100452737 | 93.09% | 187024703 | 54.60% | 84975617 |
| 440 | 96805206 | 92.74% | 179556773 | 65.90% | 61168508 |
| 441 | 98930632 | 93.51% | 185023729 | 58.70% | 76505368 |
| 442 | 96017904 | 92.77% | 178147884 | 58.20% | 74503587 |
| 443 | 90936815 | 93.73% | 170469363 | 58.90% | 70128609 |
| 444 | 89671949 | 92.11% | 165189706 | 63.80% | 59734038 |
| 445 | 93655808 | 93.56% | 175242283 | 56.20% | 76800194 |
| 446 | 92481706 | 93.39% | 172736906 | 60.80% | 67797725 |
| 447 | 92242548 | 93.29% | 172115150 | 58.20% | 71974570 |
| 448 | 98958108 | 92.46% | 183000445 | 54.90% | 82605208 |
| 449 | 89329760 | 93.54% | 167121757 | 57.40% | 71144290 |
| 450 | 92546113 | 92.53% | 171258949 | 52.70% | 80939003 |
| 451 | 95495914 | 90.44% | 172734328 | 64.70% | 60986570 |
| 452 | 90617866 | 91.86% | 166476607 | 56.70% | 72097969 |
| 453 | 85249239 | 92.77% | 158175937 | 50% | 79019520 |
| 454 | 96473089 | 93.21% | 179838851 | 61.30% | 69667130 |
| 455 | 87622388 | 93.39% | 163658186 | 52.40% | 77843675 |
| 456 | 106753538 | 95.40% | 203694475 | 53.80% | 94184243 |
| 457 | 97876344 | 94.93% | 185836410 | 59.10% | 75983609 |
| 458 | 96724752 | 94.96% | 183708439 | 56.70% | 79591673 |
| 459 | 98803703 | 94.37% | 186480153 | 56.10% | 81804328 |
| 460 | 93016321 | 94.98% | 176700272 | 54% | 81212759 |
| 461 | 92084819 | 94.83% | 174656278 | 55.80% | 77212005 |
| 462 | 95001560 | 93.67% | 177982126 | 57.20% | 76218237 |
| 463 | 106809646 | 94.65% | 202187866 | 57.80% | 85413716 |
| 464 | 87839255 | 94.12% | 165355226 | 52.80% | 77967579 |
| 465 | 105811413 | 94.38% | 199739189 | 56.70% | 86405329 |
| 466 | 95541031 | 91.59% | 175007190 | 69.10% | 54148777 |
| 467 | 100430849 | 94.49% | 189789449 | 58.90% | 77935513 |
| 468 | 105982368 | 93.82% | 198873360 | 56.20% | 87132432 |
| 469 | 101514895 | 94.94% | 192751591 | 50.30% | 95763249 |
| 470 | 94660040 | 89.61% | 169654972 | 59.10% | 69446557 |
| 471 | 101901254 | 95.58% | 194798665 | 54.90% | 87802945 |
| 472 | 94699721 | 94.57% | 179109617 | 57.20% | 76588848 |
| 473 | 105237557 | 94.78% | 199487211 | 54.10% | 91466671 |
| 474 | 95567120 | 96.24% | 183956252 | 62.70% | 68536981 |
| 475 | 94395254 | 96.17% | 181562730 | 58.70% | 74947279 |
| 476 | 98761952 | 94.66% | 186975979 | 58.10% | 78341010 |
| 477 | 92467802 | 94.89% | 175485793 | 59.10% | 71696131 |
| 478 | 96982315 | 94.40% | 183094897 | 61.40% | 70687779 |
| 479 | 101831651 | 94.93% | 193331574 | 53.20% | 90575634 |
| 480 | 97300773 | 95.01% | 184887565 | 52.60% | 87728858 |
| 481 | 96666603 | 93.43% | 180640088 | 63.40% | 66139023 |
| 482 | 102251855 | 95.72% | 195760752 | 58.30% | 81682880 |
| 483 | 97637176 | 94.63% | 184781648 | 52.90% | 86991385 |
| 484 | 101283273 | 93.99% | 190390008 | 57.20% | 81456991 |
| 485 | 101091302 | 94.91% | 191881678 | 57.90% | 80701975 |
| 486 | 93013111 | 94.11% | 175065722 | 59.80% | 70354519 |
| 487 | 105604071 | 94.43% | 199445496 | 54.20% | 91328980 |
| 488 | 93988400 | 93.25% | 175281809 | 64.30% | 62488580 |
| 489 | 99689678 | 94.93% | 189267116 | 59.30% | 77096563 |
| 490 | 103505998 | 95.05% | 196756837 | 58.80% | 80978414 |
| 491 | 97155437 | 94.88% | 184359341 | 61% | 71943927 |
| 492 | 97162311 | 94.64% | 183899317 | 64.20% | 65825221 |
| 493 | 97734107 | 94.45% | 184624265 | 62.60% | 69118740 |
| 494 | 97047216 | 94.80% | 183998443 | 63.20% | 67785009 |
| 495 | 97440239 | 94.48% | 184125148 | 54.40% | 84029016 |
| 496 | 94981444 | 94.62% | 179740485 | 62.30% | 67796232 |
| 497 | 90898018 | 93.98% | 170847926 | 62.10% | 64797984 |
| 498 | 98192837 | 95.43% | 187415265 | 57.50% | 79655358 |
| 499 | 94635900 | 95.09% | 179972994 | 57% | 77352751 |
| 500 | 96304744 | 95.18% | 183331357 | 58.10% | 76877869 |
| 501 | 99152855 | 94.85% | 188094606 | 52.80% | 88830603 |
| 502 | 87912768 | 94.13% | 165498166 | 57.90% | 69747262 |
| 503 | 102072863 | 94.93% | 193786918 | 58.80% | 79862675 |
| 504 | 95626828 | 94.70% | 181125533 | 62.10% | 68618652 |
| 505 | 95462159 | 95.32% | 181984022 | 58.90% | 74786308 |
| 506 | 86963075 | 94.99% | 165212291 | 59.70% | 66591725 |
| 507 | 102034874 | 94.81% | 193475179 | 62.70% | 72089086 |
| 508 | 93891414 | 94.86% | 178132723 | 61.10% | 69241995 |
| 509 | 95377432 | 94.81% | 180862233 | 61.40% | 69852609 |
| 510 | 105657361 | 94.56% | 199815540 | 59.40% | 81070642 |
| 511 | 94505366 | 95.15% | 179845898 | 54.60% | 81647583 |
| 512 | 93503827 | 95.19% | 178005941 | 92.80% | 12853785 |
| 513 | 97584136 | 94.64% | 184705043 | 55.40% | 82434795 |
| 514 | 94742267 | 94.40% | 178876936 | 57.80% | 75464728 |
| 515 | 96662924 | 95.12% | 183899328 | 60.60% | 72487446 |
| 516 | 95153939 | 94.63% | 180092578 | 60.40% | 71351472 |
| 517 | 94091898 | 93.90% | 176704699 | 62.10% | 66893054 |
| 518 | 92269025 | 94.15% | 173743946 | 59.40% | 70526796 |
| 519 | 94381366 | 94.39% | 178175304 | 56.20% | 77967554 |
| 520 | 95654141 | 95.06% | 181862423 | 58% | 76350924 |
| 521 | 97093342 | 94.96% | 184396026 | 58.60% | 76254633 |
| 522 | 92388585 | 95.13% | 175771350 | 58.80% | 72358164 |
| 523 | 92060426 | 94.69% | 174350333 | 58% | 73313254 |
| 524 | 109881129 | 94.29% | 207219855 | 57.20% | 88731020 |
| 525 | 101050070 | 95.68% | 193365348 | 59% | 79184265 |
| 526 | 96318962 | 94.25% | 181563940 | 58.90% | 74608502 |
| 527 | 91688583 | 94.76% | 173766923 | 58.80% | 71569344 |
| 528 | 98328743 | 95.19% | 187205952 | 58.30% | 78136105 |
| 529 | 121446366 | 94.15% | 228685610 | 57.70% | 96642845 |
| 530 | 100138016 | 94.31% | 188880646 | 61.80% | 72140808 |
| 531 | 94415088 | 95.10% | 179586762 | 57.30% | 76701379 |
| 532 | 102141157 | 95.18% | 194426179 | 55.30% | 86963987 |
| 533 | 97730240 | 95.01% | 185713182 | 54.70% | 84165544 |
| 534 | 96131064 | 95.57% | 183745808 | 55.20% | 82243616 |
| 535 | 95363612 | 94.02% | 179323841 | 59% | 73469130 |
| 536 | 102903390 | 94.13% | 193719440 | 61.20% | 75199767 |
| 537 | 100346986 | 92.46% | 185567738 | 62.10% | 70414077 |
| 538 | 100722120 | 94.73% | 190831813 | 57.70% | 80788804 |
| 539 | 95881211 | 94.30% | 180836733 | 60.70% | 71070593 |
| 540 | 95567678 | 95.28% | 182111290 | 52.60% | 86301224 |
| 541 | 97359400 | 95.05% | 185084691 | 59.30% | 75373328 |
| 542 | 97578199 | 94.91% | 185213389 | 61.30% | 71606892 |
| 543 | 96910187 | 95.58% | 185246359 | 53.70% | 85782395 |
| 544 | 112780954 | 95.18% | 214694984 | 52.20% | 102724183 |
| 545 | 93275663 | 95.35% | 177880631 | 56.20% | 77840263 |
| 546 | 83354267 | 94.64% | 157767241 | 50.80% | 77638649 |
| 547 | 90439586 | 95.09% | 171994479 | 59.60% | 69448972 |
| 548 | 103168200 | 93.69% | 193320598 | 52.20% | 92492657 |
| 549 | 98614058 | 95.08% | 187530774 | 52.90% | 88392266 |
| 550 | 98772349 | 94.43% | 186545174 | 51.80% | 89907393 |
| 551 | 96029438 | 91.99% | 176675593 | 58.20% | 73793839 |
| 552 | 94987817 | 94.13% | 178823254 | 55.70% | 79172902 |
| 553 | 98177356 | 95.20% | 186920472 | 52.80% | 88148676 |
| 554 | 94374784 | 94.06% | 177536940 | 56.20% | 77690572 |
| 555 | 99184295 | 94.78% | 188008498 | 61.80% | 71824464 |
| 556 | 96475347 | 93.23% | 179896780 | 56.40% | 78377785 |
| 557 | 95325137 | 94.82% | 180772579 | 55.80% | 79913858 |
| 558 | 99134429 | 96.11% | 190559341 | 62.80% | 70917976 |
| 559 | 95543580 | 94.65% | 180869530 | 57.70% | 76457595 |
| 560 | 101595656 | 95.06% | 193147151 | 55.40% | 86101711 |
| 561 | 91435512 | 94.70% | 173172445 | 55% | 77845967 |
| 562 | 95844954 | 93.06% | 178382933 | 60% | 71414095 |
| 563 | 93400224 | 94.81% | 177107821 | 55.50% | 78810835 |
| 564 | 97693345 | 93.78% | 183240742 | 55.80% | 81060064 |
| 565 | 111457841 | 94.01% | 209556407 | 54.40% | 95599806 |
| 566 | 100924314 | 94.69% | 191128298 | 60.80% | 74961152 |
| 567 | 95146106 | 94.99% | 180753766 | 60% | 72328773 |
| 568 | 92308177 | 94.52% | 174506357 | 60.40% | 69065020 |
| 569 | 93055956 | 94.71% | 176261034 | 53.60% | 81708334 |
| 570 | 94736443 | 94.54% | 179120753 | 59.50% | 72515103 |
| 571 | 101645086 | 94.82% | 192751968 | 59.50% | 78029324 |
| 572 | 95245554 | 93.82% | 178711513 | 58.70% | 73752412 |
| 573 | 94462242 | 94.09% | 177753492 | 58.60% | 73591688 |
| 574 | 94920698 | 94.50% | 179406360 | 57.90% | 75451736 |
| 575 | 93789701 | 95.31% | 178776881 | 57.80% | 75530134 |

**Supplementary Table 2:**
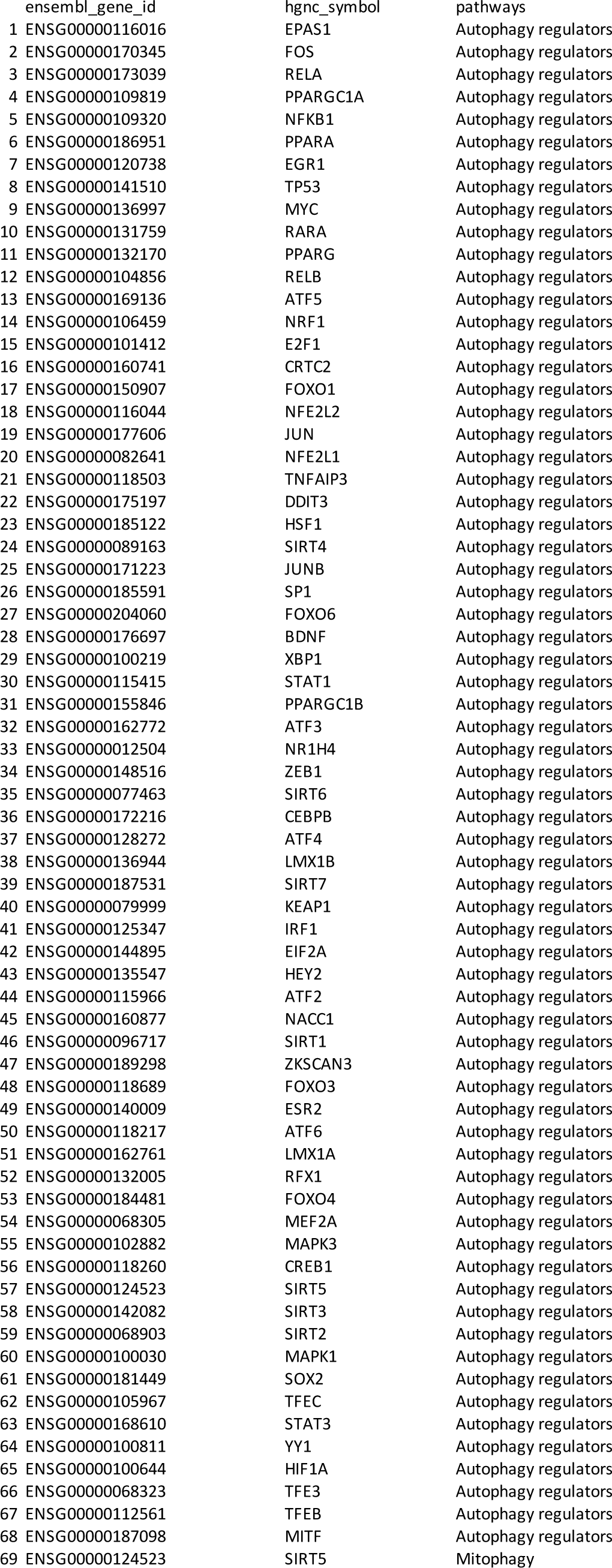
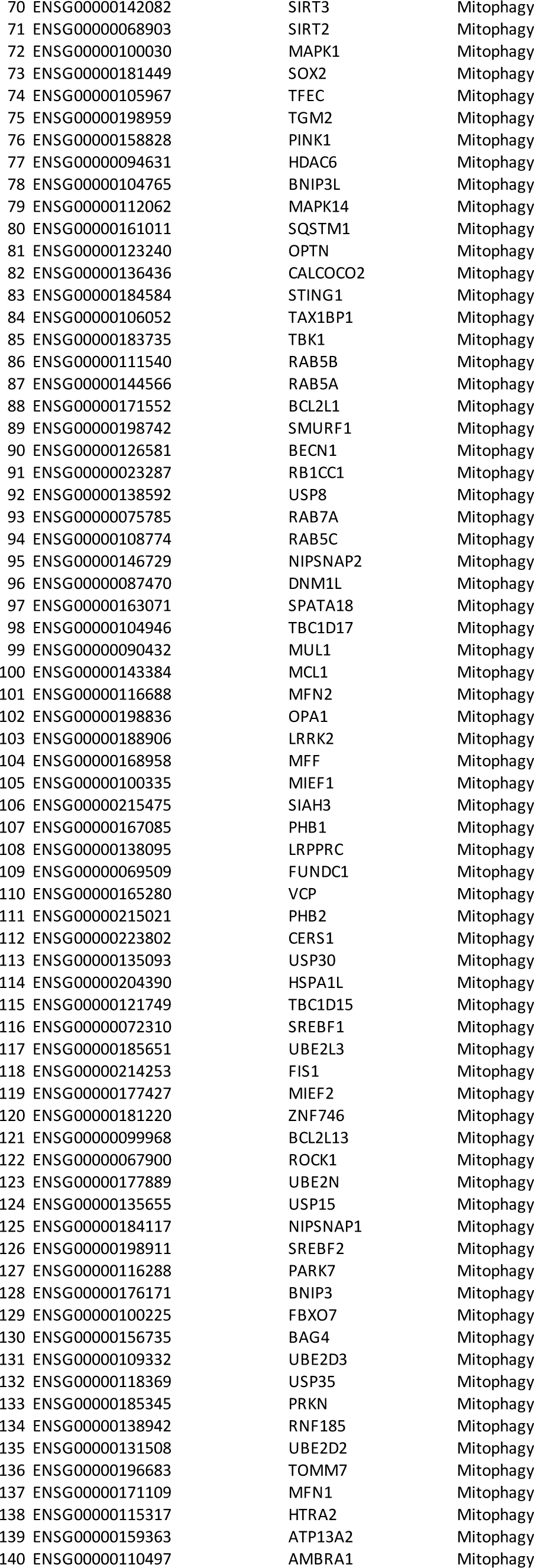
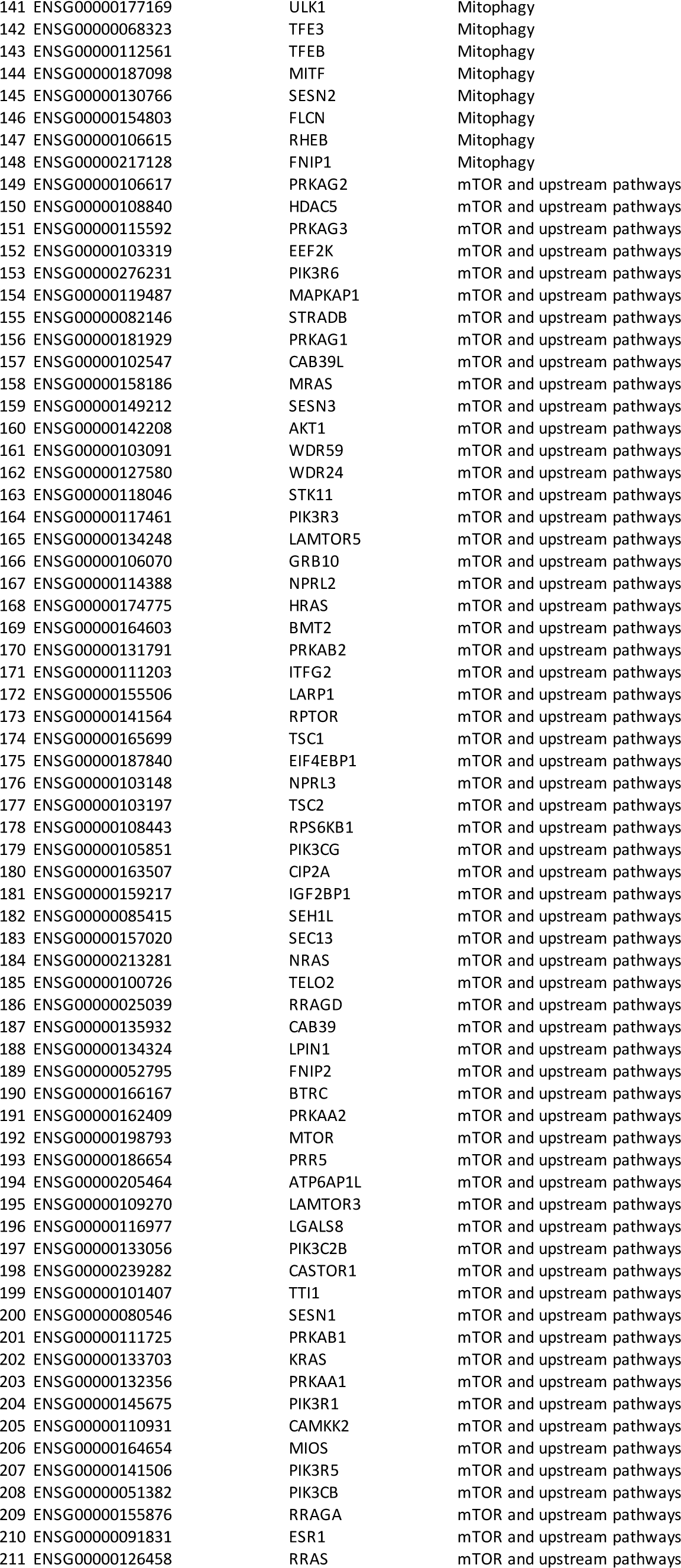
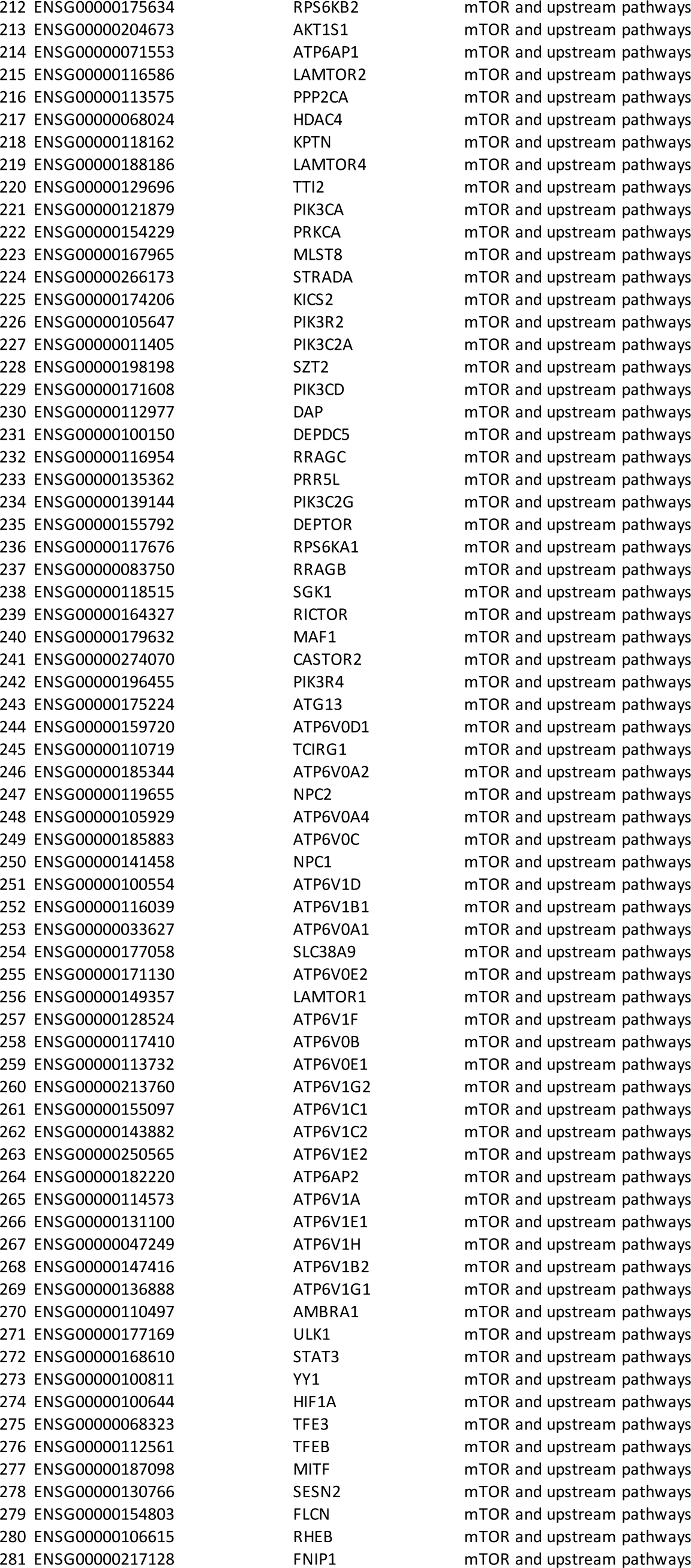
List of 281 autophagy genes with ENSG identifiers.

**Supplementary Tables 3-8:**
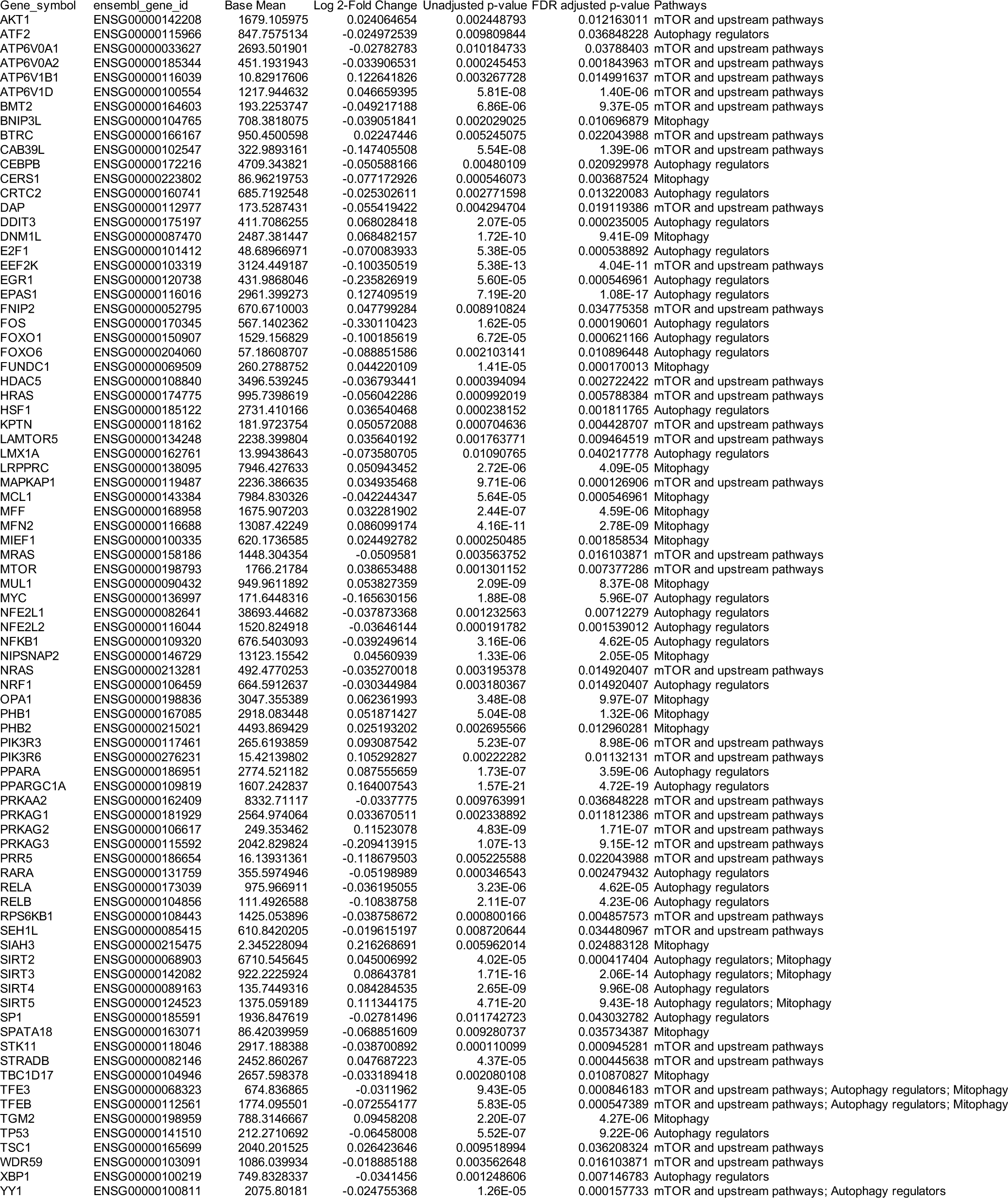
Listings of Ensembl gene (ENSG) associations identified by negative binomial regression for each trait (table 3: Max OXPHOS, table 4: VO_2_ peak, table 5: 400-meter Walk Speed, table 6: Leg Strength, table 7: Leg Power, table 8: Thigh Muscle Volume).

**Supplementary Table 4.**
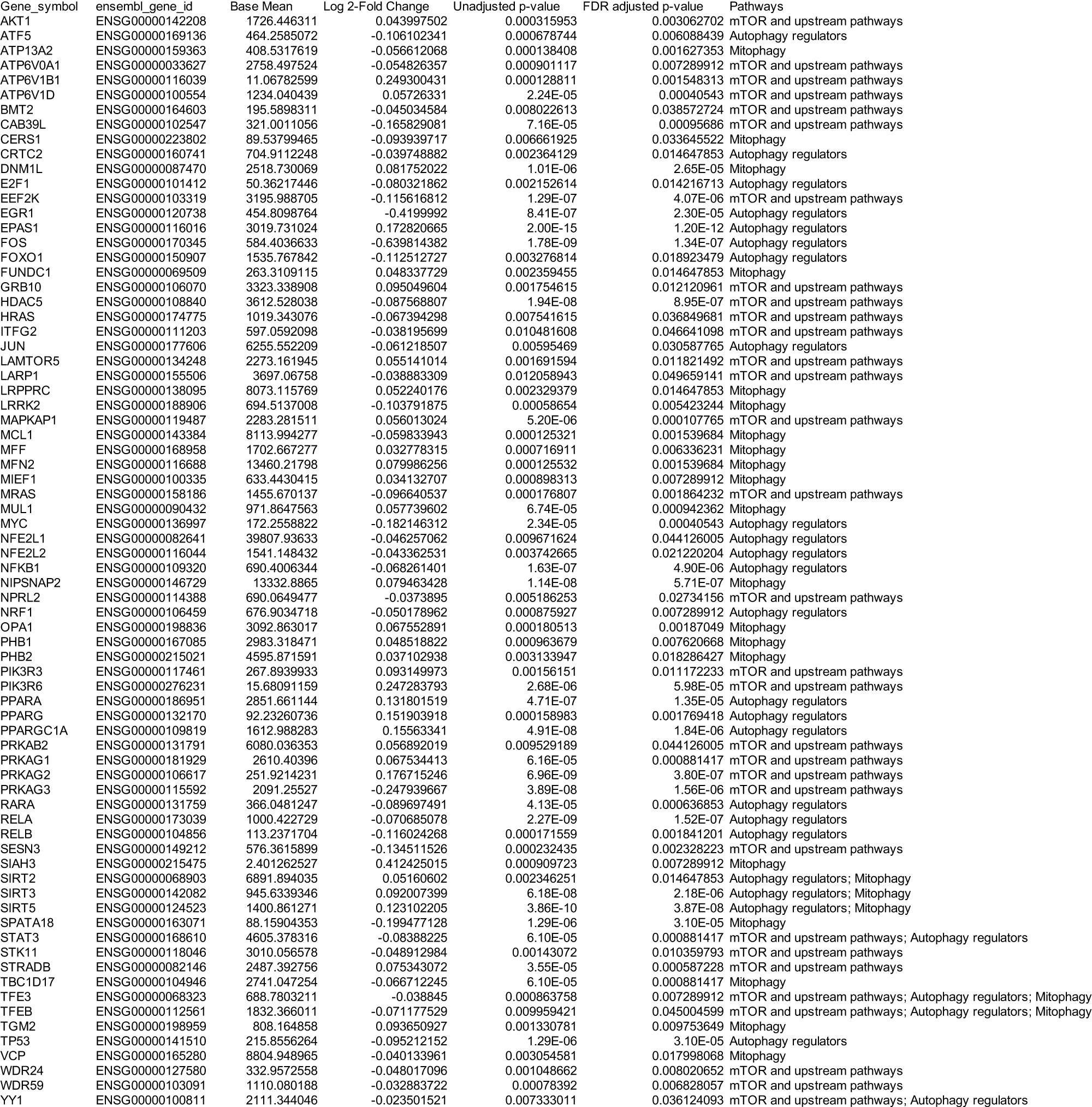

**Supplementary Table 5.**
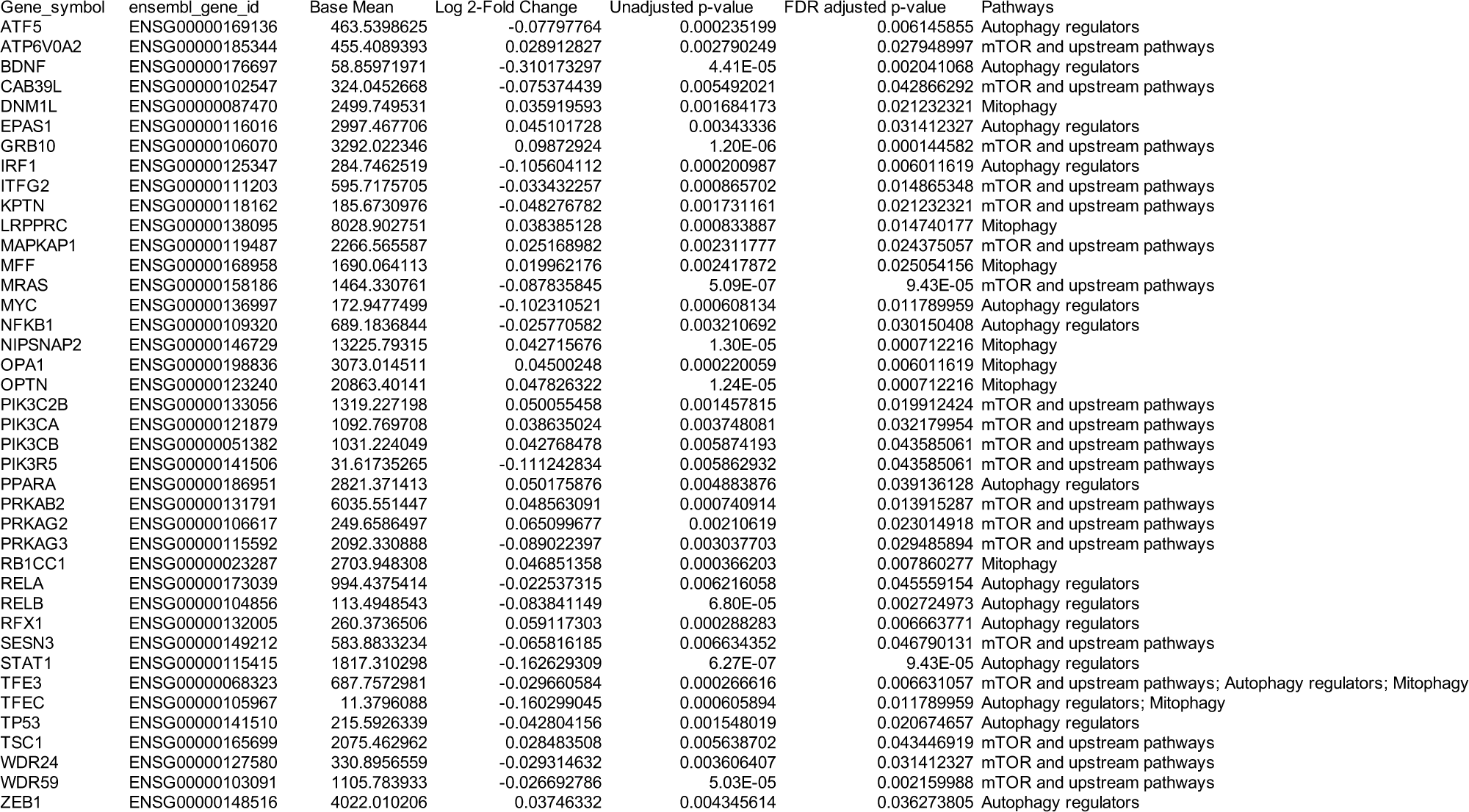

**Supplementary Table 6.**
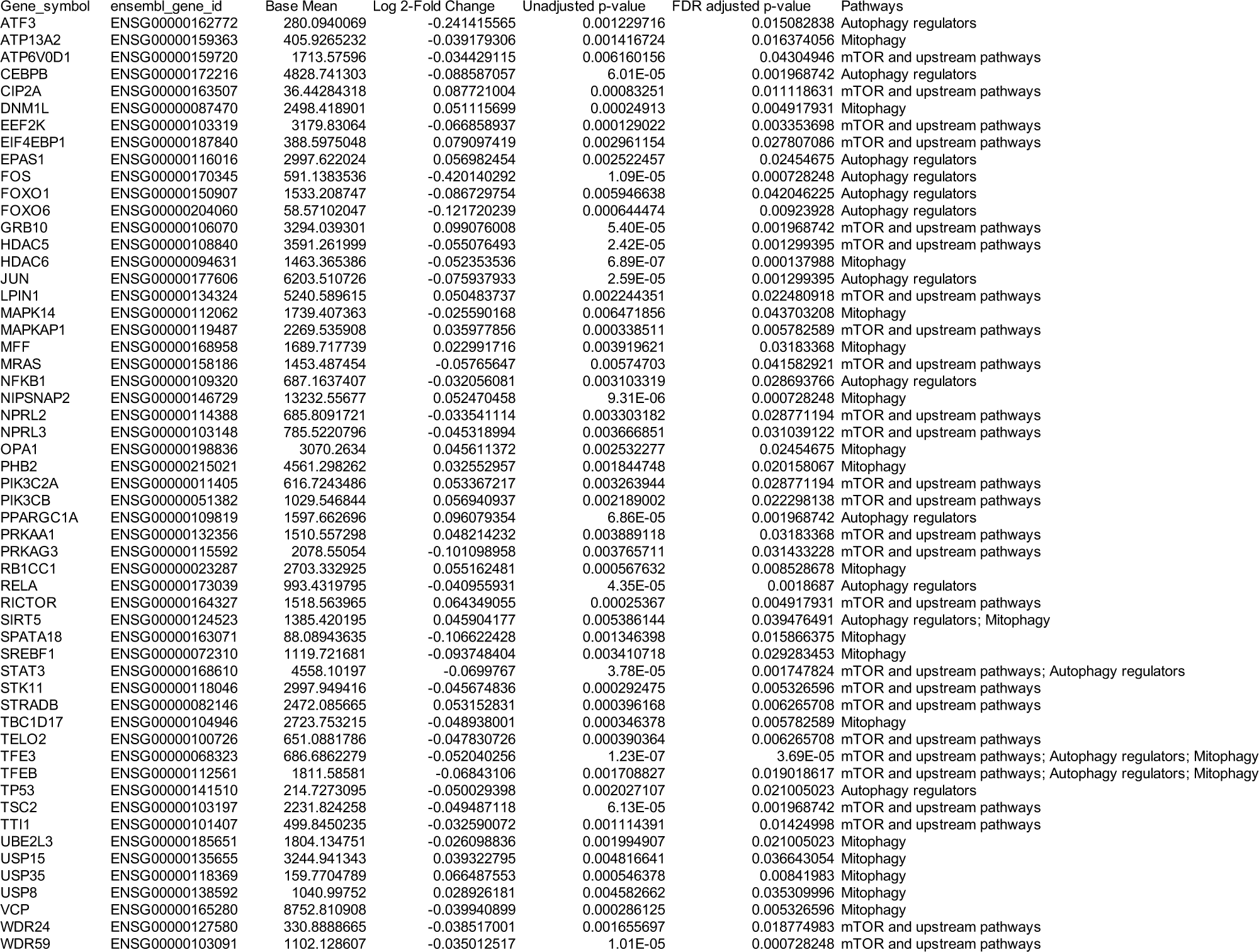

**Supplementary Table 7.**
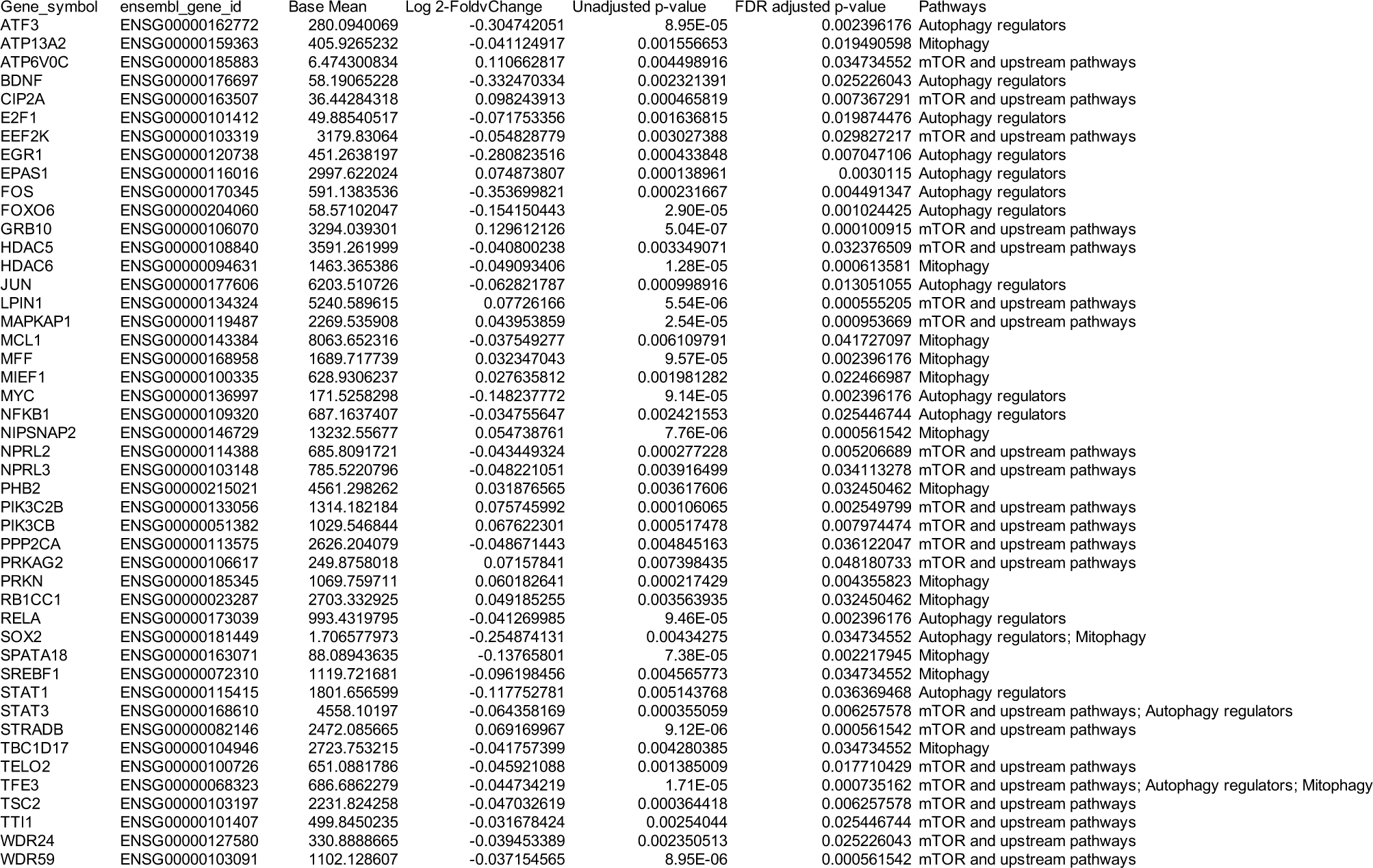

**Supplementary Table 8.**
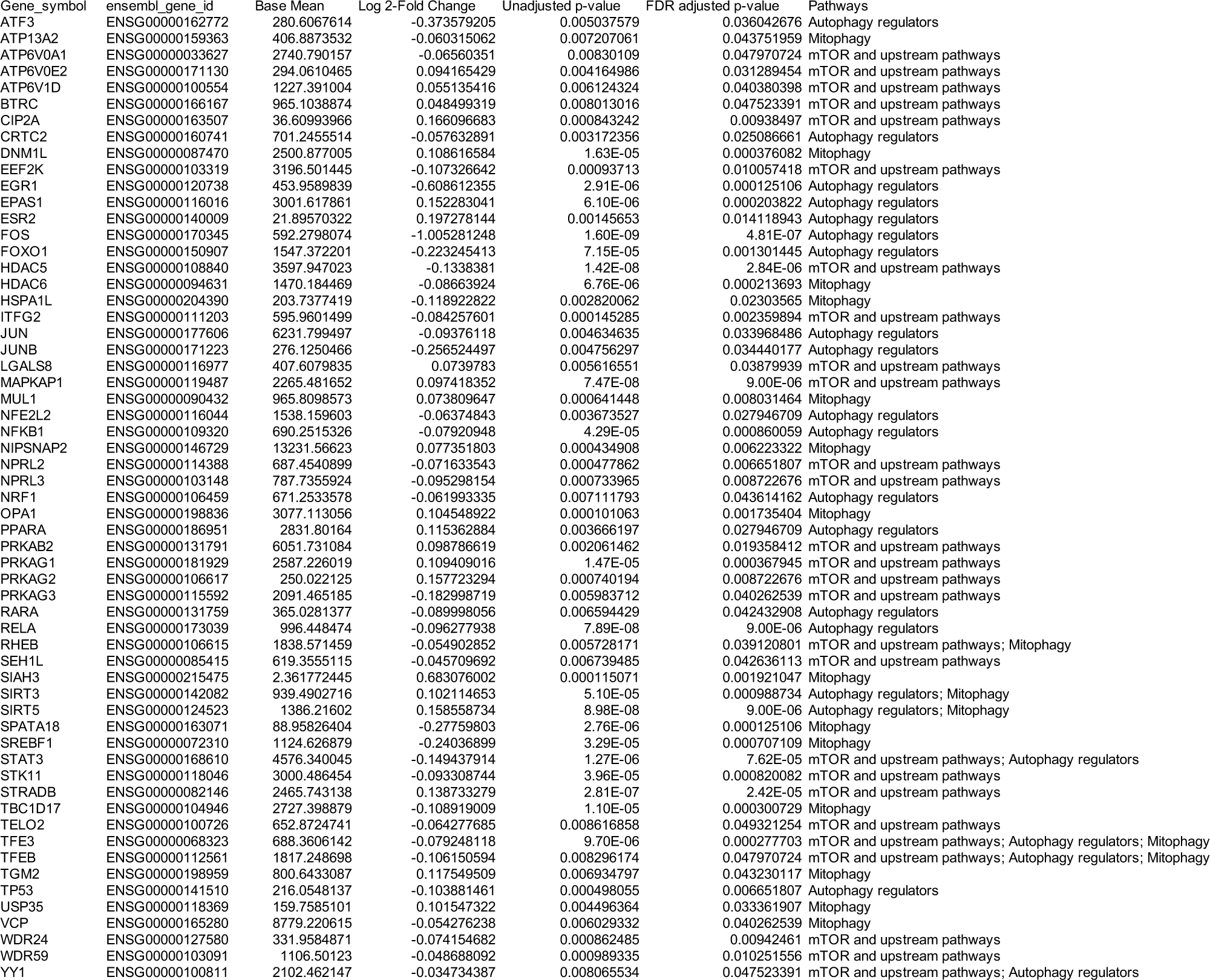

| Gene_symbol | ensembl_gene_id | Base Mean | Log 2-Fold Change | Unadjusted p-value | FDR adjusted p-value | Pathways |
| --- | --- | --- | --- | --- | --- | --- |
| ATF3 | ENSG00000162772 | 280.6067614 | -0.373579205 | 0.005037579 | 0.036042676 | Autophagy regulators |
| ATP13A2 | ENSG00000159363 | 406.8873532 | -0.060315062 | 0.007207061 | 0.043751959 | Mitophagy |
| ATP6V0A1 | ENSG00000033627 | 2740.790157 | -0.06560351 | 0.00830109 | 0.047970724 | mTOR and upstream pathways |
| ATP6V0E2 | ENSG00000171130 | 294.0610465 | 0.094165429 | 0.004164986 | 0.031289454 | mTOR and upstream pathways |
| ATP6V1D | ENSG00000100554 | 1227.391004 | 0.055135416 | 0.006124324 | 0.040380398 | mTOR and upstream pathways |
| BTRC | ENSG00000166167 | 965.1038874 | 0.048499319 | 0.008013016 | 0.047523391 | mTOR and upstream pathways |
| CIP2A | ENSG00000163507 | 36.60993966 | 0.166096683 | 0.000843242 | 0.00938497 | mTOR and upstream pathways |
| CRTC2 | ENSG00000160741 | 701.2455514 | -0.057632891 | 0.003172356 | 0.025086661 | Autophagy regulators |
| DNM1L | ENSG00000087470 | 2500.877005 | 0.108616584 | 1.63E-05 | 0.000376082 | Mitophagy |
| EEF2K | ENSG00000103319 | 3196.501445 | -0.107326642 | 0.00093713 | 0.010057418 | mTOR and upstream pathways |
| EGR1 | ENSG00000120738 | 453.9589839 | -0.608612355 | 2.91E-06 | 0.000125106 | Autophagy regulators |
| EPAS1 | ENSG00000116016 | 3001.617861 | 0.152283041 | 6.10E-06 | 0.000203822 | Autophagy regulators |
| ESR2 | ENSG00000140009 | 21.89570322 | 0.197278144 | 0.00145653 | 0.014118943 | Autophagy regulators |
| FOS | ENSG00000170345 | 592.2798074 | -1.005281248 | 1.60E-09 | 4.81E-07 | Autophagy regulators |
| FOXO1 | ENSG00000150907 | 1547.372201 | -0.223245413 | 7.15E-05 | 0.001301445 | Autophagy regulators |
| HDAC5 | ENSG00000108840 | 3597.947023 | -0.1338381 | 1.42E-08 | 2.84E-06 | mTOR and upstream pathways |
| HDAC6 | ENSG00000094631 | 1470.184469 | -0.08663924 | 6.76E-06 | 0.000213693 | Mitophagy |
| HSPA1L | ENSG00000204390 | 203.7377419 | -0.118922822 | 0.002820062 | 0.02303565 | Mitophagy |
| ITFG2 | ENSG00000111203 | 595.9601499 | -0.084257601 | 0.000145285 | 0.002359894 | mTOR and upstream pathways |
| JUN | ENSG00000177606 | 6231.799497 | -0.09376118 | 0.004634635 | 0.033968486 | Autophagy regulators |
| JUNB | ENSG00000171223 | 276.1250466 | -0.256524497 | 0.004756297 | 0.034440177 | Autophagy regulators |
| LGALS8 | ENSG00000116977 | 407.6079835 | 0.0739783 | 0.005616551 | 0.03879939 | mTOR and upstream pathways |
| MAPKAP1 | ENSG00000119487 | 2265.481652 | 0.097418352 | 7.47E-08 | 9.00E-06 | mTOR and upstream pathways |
| MUL1 | ENSG00000090432 | 965.8098573 | 0.073809647 | 0.000641448 | 0.008031464 | Mitophagy |
| NFE2L2 | ENSG00000116044 | 1538.159603 | -0.06374843 | 0.003673527 | 0.027946709 | Autophagy regulators |
| NFKB1 | ENSG00000109320 | 690.2515326 | -0.07920948 | 4.29E-05 | 0.000860059 | Autophagy regulators |
| NIPSNAP2 | ENSG00000146729 | 13231.56623 | 0.077351803 | 0.000434908 | 0.006223322 | Mitophagy |
| NPRL2 | ENSG00000114388 | 687.4540899 | -0.071633543 | 0.000477862 | 0.006651807 | mTOR and upstream pathways |
| NPRL3 | ENSG00000103148 | 787.7355924 | -0.095298154 | 0.000733965 | 0.008722676 | mTOR and upstream pathways |
| NRF1 | ENSG00000106459 | 671.2533578 | -0.061993335 | 0.007111793 | 0.043614162 | Autophagy regulators |
| OPA1 | ENSG00000198836 | 3077.113056 | 0.104548922 | 0.000101063 | 0.001735404 | Mitophagy |
| PPARA | ENSG00000186951 | 2831.80164 | 0.115362884 | 0.003666197 | 0.027946709 | Autophagy regulators |
| PRKAB2 | ENSG00000131791 | 6051.731084 | 0.098786619 | 0.002061462 | 0.019358412 | mTOR and upstream pathways |
| PRKAG1 | ENSG00000181929 | 2587.226019 | 0.109409016 | 1.47E-05 | 0.000367945 | mTOR and upstream pathways |
| PRKAG2 | ENSG00000106617 | 250.022125 | 0.157723294 | 0.000740194 | 0.008722676 | mTOR and upstream pathways |
| PRKAG3 | ENSG00000115592 | 2091.465185 | -0.182998719 | 0.005983712 | 0.040262539 | mTOR and upstream pathways |
| RARA | ENSG00000131759 | 365.0281377 | -0.089998056 | 0.006594429 | 0.042432908 | Autophagy regulators |
| RELA | ENSG00000173039 | 996.448474 | -0.096277938 | 7.89E-08 | 9.00E-06 | Autophagy regulators |
| RHEB | ENSG00000106615 | 1838.571459 | -0.054902852 | 0.005728171 | 0.039120801 | mTOR and upstream pathways; Mitophagy |
| SEH1L | ENSG00000085415 | 619.3555115 | -0.045709692 | 0.006739485 | 0.042636113 | mTOR and upstream pathways |
| SIAH3 | ENSG00000215475 | 2.361772445 | 0.683076002 | 0.000115071 | 0.001921047 | Mitophagy |
| SIRT3 | ENSG00000142082 | 939.4902716 | 0.102114653 | 5.10E-05 | 0.000988734 | Autophagy regulators; Mitophagy |
| SIRT5 | ENSG00000124523 | 1386.21602 | 0.158558734 | 8.98E-08 | 9.00E-06 | Autophagy regulators; Mitophagy |
| SPATA18 | ENSG00000163071 | 88.95826404 | -0.27759803 | 2.76E-06 | 0.000125106 | Mitophagy |
| SREBF1 | ENSG00000072310 | 1124.626879 | -0.24036899 | 3.29E-05 | 0.000707109 | Mitophagy |
| STAT3 | ENSG00000168610 | 4576.340045 | -0.149437914 | 1.27E-06 | 7.62E-05 | mTOR and upstream pathways; Autophagy regulators |
| STK11 | ENSG00000118046 | 3000.486454 | -0.093308744 | 3.96E-05 | 0.000820082 | mTOR and upstream pathways |
| STRADB | ENSG00000082146 | 2465.743138 | 0.138733279 | 2.81E-07 | 2.42E-05 | mTOR and upstream pathways |
| TBC1D17 | ENSG00000104946 | 2727.398879 | -0.108919009 | 1.10E-05 | 0.000300729 | Mitophagy |
| TELO2 | ENSG00000100726 | 652.8724741 | -0.064277685 | 0.008616858 | 0.049321254 | mTOR and upstream pathways |
| TFE3 | ENSG00000068323 | 688.3606142 | -0.079248118 | 9.70E-06 | 0.000277703 | mTOR and upstream pathways; Autophagy regulators; Mitophagy |
| TFEB | ENSG00000112561 | 1817.248698 | -0.106150594 | 0.008296174 | 0.047970724 | mTOR and upstream pathways; Autophagy regulators; Mitophagy |
| TGM2 | ENSG00000198959 | 800.6433087 | 0.117549509 | 0.006934797 | 0.043230117 | Mitophagy |
| TP53 | ENSG00000141510 | 216.0548137 | -0.103881461 | 0.000498055 | 0.006651807 | Autophagy regulators |
| USP35 | ENSG00000118369 | 159.7585101 | 0.101547322 | 0.004496364 | 0.033361907 | Mitophagy |
| VCP | ENSG00000165280 | 8779.220615 | -0.054276238 | 0.006029332 | 0.040262539 | Mitophagy |
| WDR24 | ENSG00000127580 | 331.9584871 | -0.074154682 | 0.000862485 | 0.00942461 | mTOR and upstream pathways |
| WDR59 | ENSG00000103091 | 1106.50123 | -0.048688092 | 0.000989335 | 0.010251556 | mTOR and upstream pathways |
| YY1 | ENSG00000100811 | 2102.462147 | -0.034734387 | 0.008065534 | 0.047523391 | mTOR and upstream pathways; Autophagy regulators |

